# Short-term forecasting of COVID-19 in Germany and Poland during the second wave – a preregistered study

**DOI:** 10.1101/2020.12.24.20248826

**Authors:** J. Bracher, D. Wolffram, J. Deuschel, K. Görgen, J.L. Ketterer, A. Ullrich, S. Abbott, M.V. Barbarossa, D. Bertsimas, S. Bhatia, M. Bodych, N.I. Bosse, J.P. Burgard, L. Castro, G. Fairchild, J. Fuhrmann, S. Funk, K. Gogolewski, Q. Gu, S. Heyder, T. Hotz, Y. Kheifetz, H. Kirsten, T. Krueger, E. Krymova, M.L. Li, J.H. Meinke, I.J. Michaud, K. Niedzielewski, T. Ożański, F. Rakowski, M. Scholz, S. Soni, A. Srivastava, J. Zieliński, D. Zou, T. Gneiting, M. Schienle

## Abstract

We report insights from ten weeks of collaborative COVID-19 forecasting for Germany and Poland (12 October – 19 December 2020). The study period covers the onset of the second wave in both countries, with tightening non-pharmaceutical interventions (NPIs) and subsequently a decay (Poland) or plateau and renewed increase (Germany) in reported cases. Thirteen independent teams provided probabilistic real-time forecasts of COVID-19 cases and deaths. These were reported for lead times of one to four weeks, with evaluation focused on one- and two-week horizons, which are less affected by changing NPIs. Heterogeneity between forecasts was considerable both in terms of point predictions and forecast spread. Ensemble forecasts showed good relative performance, in particular in terms of coverage, but did not clearly dominate single-model predictions. The study was preregistered and will be followed up in future phases of the pandemic.

## 1 Introduction

Forecasting is one of the key purposes of epidemic modelling, and despite being related to the understanding of underlying mechanisms, it is a conceptually distinct task (Keeling and Rohani, 2008). Accurate disease forecasts can improve situational awareness of decision makers and facilitate tasks such as resource allocation or planning of vaccine trials (Dean et al., 2020). During the COVID-19 pandemic, there has been a major surge in research activity on epidemic forecasting with a plethora of approaches being pursued. Contributions vary greatly in terms of purpose, forecast targets, methods, and evaluation criteria. An important distinction is between *longer-term scenario* or *what-if projections* and *short-term forecasts* (Reich and Rivers, 2020). The former attempt to discern the consequences of hypothetical scenarios and typically cannot be evaluated directly using subsequently observed data. The latter, which are the focus of this work, quantitatively describe expectations and uncertainties in the short run. They refer to quantities expected to be largely unaffected by yet unknown changes in public health interventions. This makes them particularly suitable to assess the predictive power of computational models, a need repeatedly expressed during the pandemic (Nature Publishing Group, 2020).

In this work we present results and takeaways from a collaborative and prospective short-term COVID-19 forecasting project in Germany and Poland. The evaluation period extends from 12 October 2020 (first forecasts issued) to 19 December 2020 (last observations made), thus covering the onset of the second epidemic wave in both countries. We gathered a total of 13 modelling teams from Germany, Poland, Switzerland, the United Kingdom and the United States to generate forecasts of confirmed cases and deaths in a standardized and thus comparable manner. These are publicly available in an online repository (https://github.com/KITmetricslab/covid19-forecast-hub-de) called the *German and Polish COVID-19 Forecast Hub* and can be explored interactively in a dashboard (https://kitmetricslab.github.io/forecasthub). On 8 October 2020, we deposited a study protocol (Bracher et al., 2020b) at the registry of the Open Science Foundation (OSF), predefining the study period and procedures for a prospective forecast evaluation study. Here we report on results from this effort, addressing in particular the following questions:

- At which forecast horizons can one expect to obtain reliable forecasts for various targets?
- Are the forecasts calibrated, i.e. are they able to accurately quantify their own uncertainty?
- How good is the agreement between different forecast methods?
- Are there prediction approaches which prove to be particularly reliable?
- Can combined ensemble forecasts lead to improved performance?

The study period is marked by overall strong virus circulation and changes in intervention measures and testing strategies. This makes for a situation in which reliable short-term predictions are both particularly useful and particularly challenging to produce. Conclusions from ten weeks of real-time forecasting are necessarily preliminary, but we hope to contribute to an ongoing exchange on best practices in the field. Our study will be followed up until at least March 2021 and may be extended beyond.

The project follows several principles which we consider key for a rigorous assessment of forecasting methods. Firstly, forecasts are made in real time, as retrospective forecasting often leads to overly optimistic conclusions about performance. Real-time forecasting poses many specific challenges (Desai et al., 2019), including noisy or delayed data, incomplete knowledge on testing and interventions as well as time pressure. Even if these are mimicked in retrospective studies, some benefit of hindsight remains. Secondly, in a pandemic situation with presumably low predictability we consider it of central importance to explicitly quantify forecast uncertainty. Forecasts should thus be probabilistic rather than limited to point forecasts (Held et al., 2017; Funk et al., 2019). Lastly, forecast studies are most informative if they involve statistically sound comparisons between multiple independently run forecast methods (Viboud and Vespignani, 2019). We therefore aimed for a body of standardized, comparable and uniformly formatted short-term forecasts. Such collaborative efforts have led to important advances in short-term disease forecasting prior to the pandemic (Viboud et al., 2018; Del Valle et al., 2018; Johansson et al., 2019; Reich et al., 2019a). Notably, they have provided evidence that ensemble forecasts combining various independent predictions can lead to improved performance, similar to what has been observed in weather prediction (Gneiting and Raftery, 2005).

The German and Polish Forecast Hub project also aims to provide a platform for exchange between research teams from both countries and beyond. To this end, regular video conferences with presentations and discussions on forecast methodologies were organized. Moreover, the Forecast Hub Team provided feedback on performance in order to facilitate model revisions and forecast improvement.

The German and Polish COVID-19 Forecast Hub is run in close exchange with the US COVID-19 Forecast Hub (Ray et al. 2020; COVID-19 Forecast Hub Team 2020) and aims for compatibility with the short-term forecasts assembled there. Consequently, many formal aspects presented in Section 2 are shared between the two projects. However, we faced a number of distinct challenges, including rapid changes in non-pharmaceutical interventions, the use of different truth data sources by different teams and a smaller number of contributing teams. Close links moreover exist to a similar effort in the United Kingdom (Funk et al., 2020). Other conceptually related works on short-term forecasting or baseline projections include those by the Austrian COVID-19 Forecast Consortium (Bicher et al., 2020) and the European Centre for Disease Prevention and Control (ECDC; 2020a; 2020c). In a German context, various nowcasting efforts exist, see e.g. Günther et al. (2020).

## 2 Formal setting

We start by laying out the formal framework of the presented collaborative forecasting study. Unless stated differently, the principles correspond to those specified in the study protocol (Bracher et al., 2020b).

### 2.1 Submission system and rhythm

All submissions were collected in a standardized format in a public repository to which teams could submit (https://github.com/KITmetricslab/covid19-forecast-hub-de). For teams running their own repositories, the Forecast Hub Team put in place software scripts to re-format forecasts and transfer them into the Hub repository. Participating teams were asked to update their forecasts on a weekly basis using data up to Monday. Submission was possible until Tuesday 3 pm Berlin/Warsaw time. Delayed submission of forecasts was possible until Wednesday, with exceptional further extensions possible in case of technical issues. Delays of submissions were documented (Supplementary Table 4).

### 2.2 Forecast targets and format

We focus on short-term forecasting of confirmed cases and deaths from COVID-19 in Germany and Poland one and two weeks ahead. Here, weeks refer to Morbidity and Mortality Weekly Report (MMWR) weeks which start on Sunday and end on Saturday, meaning that one-week-ahead forecasts were actually five days ahead, two-week ahead forecasts were twelve days ahead, etc. All targets were defined by the date of reporting to the national authorities (rather than e.g. symptom onset date). This means that modellers have to take reporting delays into account, but has the advantage that data points are usually not revised over the following days and weeks. All targets were addressed both on cumulative and weekly incident scales. Forecasts could refer to both data from the European Centre for Disease Prevention and Control (ECDC; 2020b) and Johns Hopkins University Center for Systems Science and Engineering (JHU CSSE; Dong et al. 2020). In this article we focus on the preregistered period of 12 October 2020 to 19 December 2020. Figure 1 shows the targets on an incidence scale for the two countries, with the study period highlighted. We also indicate the timing of changes in interventions and reporting procedures which were considered of importance for short-term forecasting.

**Figure 1:**
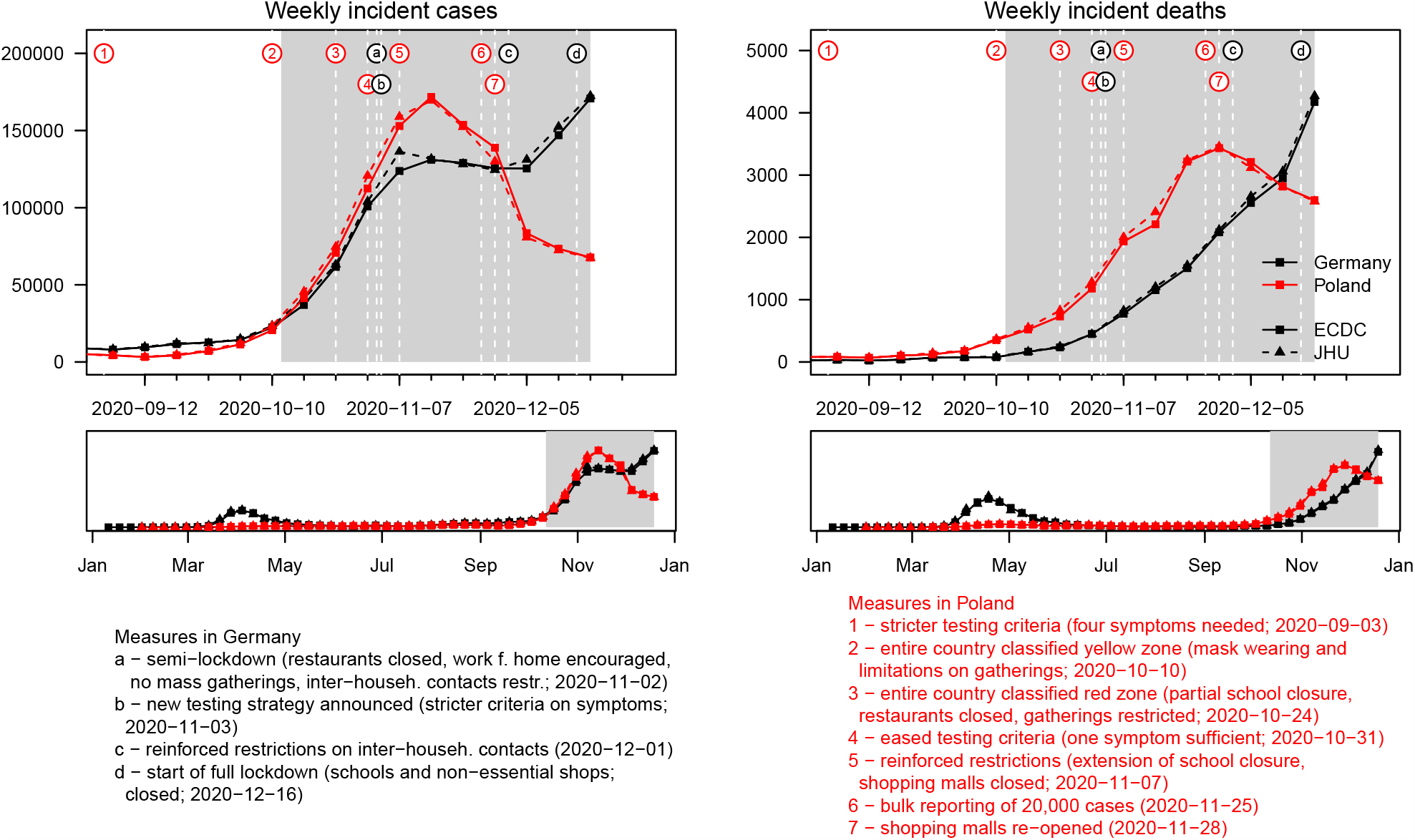
Weekly incident confirmed cases and deaths from COVID-19 in Germany and Poland according to data sets from ECDC and JHU. The study period covered in this paper is highlighted in grey. Important changes in interventions and testing are marked. Sources containing details on the listed interventions are provided in Supplementary Section C.

Note that on 14 December 2020, the ECDC data set on COVID-19 cases and deaths in daily resolution was discontinued. For the last weekly data point we therefore used data streams from Robert Koch Institute and the Polish Ministry of Health which we had previously used to obtain regional data and which up to this time had been in agreement with the ECDC data.

Most forecasters also produced and submitted three- and four-week-ahead forecasts (which were specified as targets in the study protocol). These horizons, also used in the US COVID-19 Forecast Hub (Ray et al., 2020), were originally defined for deaths. Due to their lagged nature, these were considered predictable independently of future policy or behavioural changes up to four weeks ahead; see UK Scientific Pandemic Influenza Group on Modelling (2020) for a similar argument. During the summer months, when incidence was low and intervention measures largely constant, the same horizons were introduced for cases. As the epidemic situation and intervention measures became more dynamic in autumn, it became clear that case forecasts further than two weeks (twelve days) ahead were too dependent on yet unknown interventions and the consequent changes in transmission rates. It was therefore decided to restrict the default view in the online dashboard to one- and two-week-ahead forecasts only. At the same time we continued to collect three- and four-week-ahead outputs. Most models (with the exception of `epiforecasts-EpiExpert`, `COVIDAnalytics-Delphi` and in some exceptional cases `MOCOS-agent1`) do not anticipate policy changes, so that their outputs can be seen as “baseline projections”, i.e. projections for a scenario with constant interventions. In accordance with the study protocol we also report on three- and four-week-ahead predictions, but these results have been moved to the Supplementary Material.

We emphasize the importance of quantifying the uncertainty associated with forecasts. Teams were therefore asked to report a total of 23 predictive quantiles (1%, 2.5%, 5%, 10%, …, 90%, 95%, 97.5%, 99%) in addition to their point forecasts. This motivates considering both forecasts of cumulative and incident quantities, as predictive quantiles for these generally cannot be translated from one scale to the other. Not all teams provided such probabilistic forecasts, though, and we also accepted pure point forecasts.

### 2.3 Evaluation measures

The submitted quantiles of a predictive distribution *F* define 11 central prediction intervals with nominal coverage level 1 *− α* where *α* = 0.02, 0.05, 0.10, 0.20, …, 0.90. Each of these can be evaluated using the *interval score* (Gneiting and Raftery, 2007):

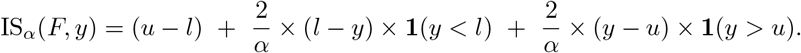

Here *u* and *l* are the lower and upper ends of the respective interval, **1** is the indicator function and *y* is the eventually observed value. The three summands can be interpreted as a measure of sharpness and penalties for under- and overprediction, respectively. The primary evaluation measure used in this study is the *weighted interval score* (WIS; Bracher et al. 2020a), which combines the absolute error (AE) of the predictive median *m* and the interval scores achieved for the eleven nominal levels. The WIS is a well-known quantile-based approximation of the continuous ranked probability score (CRPS; Gneiting and Raftery 2007) and, in the case of our 11 intervals, defined as

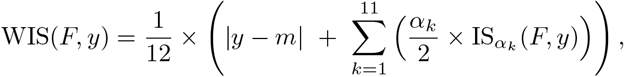

where *α*_1_ = 0.02, *α*_2_ = 0.05, *α*_3_ = 0.10, *α*_4_ = 0.20, …, *α*_11_ = 0.90. The score reflects the distance between the predictive distribution *F* and the eventually observed outcome *y*, and thus is negatively oriented, meaning that smaller values are better. As secondary measures of forecast performance we considered the absolute error of point forecasts and the empirical coverage of 50% and 95% prediction intervals. In this context we note that WIS and AE are equivalent for deterministic forecasts (i.e. forecasts concentrating all probability mass on a single value). This enables a principled comparison between probabilistic and deterministic forecasts, both of which appear in the present study.

In the evaluation we needed to account for the fact that forecasts can refer to either the ECDC or JHU data sets. We performed all forecast evaluations once using ECDC data and once using JHU data, with ECDC being our prespecified primary data source. For cumulative targets we shifted forecasts which refer to the other truth data source additively by the last observed difference. This is a pragmatic strategy to align forecasts with the last state of the respective time series.

Another difficulty in comparative forecast evaluation lies in the handling of missing forecasts. For this case (which indeed occurred for several teams) we prespecified that the missing score would be imputed with the worst (i.e. largest) score obtained by any other forecaster. In the respective summary tables any such instances are marked. All values reported are mean scores over the evaluation period, though if more than a third of the forecasts were missing we refrain from reporting.

## 3 Forecasting methods

### 3.1 Baseline forecasts

In order to put evaluation results into perspective we use three simple reference models. Note that only the first was prespecified. The two others were added later as the need for comparisons to simple, but not completely naïve, approaches was recognized. More detailed descriptions are provided in Supplementary Section B.

`KIT-baseline`: A naïve last-observation carried-forward approach (on the incidence scale) with identical variability for all forecast horizons (estimated from the last five observations). This is very similar to the *null model* used by Funk et al. (2020).

`KIT-extrapolation_baseline`: A multiplicative extrapolation based on the last two observations with uncertainty bands estimated from five preceding observations.

`KIT-time_series_baseline` An exponential smoothing model with multiplicative error terms and no seasonality as implemented in the `R` package `forecast` (Hyndman and Khandakar, 2008) and used for COVID-19 forecasting by Petropoulos and Makridakis (2020).

### 3.2 Contributed forecasts

During the evaluation period from October to December 2020, we assembled short-term predictions from a total of 14 forecast methods by 13 independent teams of researchers. Eight of these are run by teams collaborating directly with the Hub, based on models these researchers were either already running or set up specifically for the purpose of short-term forecasting. The remaining short-term forecasts were made available via dedicated online dashboards by their respective authors, often along with forecasts for other countries. With their permission, the Forecast Hub team assembled and integrated these forecasts. Table 1 provides an overview of all included models with brief descriptions and information on the handling of non-pharmaceutical interventions, testing strategies, age strata and the source used for truth data. The models span a wide range of approaches, from computationally expensive agent-based simulations to human judgement forecasts. Not all models addressed all targets and forecast horizons suggested in our project; which targets were addressed by which models can be seen from Tables 2 and 3.

**Table 1:** Forecast models contributed by independent external research teams. Abbreviations: NPI: Does the forecast model explicitly account for non-pharmaceutical interventions? Test: Does the model account for changing testing strategies? Age: Is the model age-structured? DE, PL: Are forecasts issued for Germany and Poland, respectively? Truth: Which truth data source does the model use? Pr: Are forecasts probabilistic (23 quantiles)?

| Category | Model | Description | NPI | Test | Age | DE | PL | Truth | Pr |
| --- | --- | --- | --- | --- | --- | --- | --- | --- | --- |
| Agent-based | ICM-agentModel | Agent-based model for stochastic simulations of air-borne disease spread. Agents are assigned to geographically distributed contexts. The model implements a travel module that moves agents between cities (Rakowski et al., 2010). | ✓ | ✓ | ✓ |  | ✓ | JHU | ✓ |
|  | MOCOS-agent1 | Agent based model. Continuous-time stochastic microsimulation based on census data, including contact tracing, testing and quarantine (Adamik et al., 2020). Relevant duration time distributions are based on empirical data. | ✓ | ✓ | ✓ |  | ✓ | JHU | ✓ |
| Compartment | CovidAnalytics-DELPHI<sup>1</sup> | Country-level modified SEIR model accounting for changing interventions and underdetection (Li et al., 2020). | ✓ |  |  | ✓ | ✓ | JHU | ✓ |
|  | FIAS_FZJ-Epi1Ger | Country-level deterministic model, extension of classical SEIR approach, takes explicitly into account undetected cases and reporting delays (Barbarossa et al., 2020). |  |  |  | ✓ |  | ECDC | ✓ |
|  | LeipzigIMISE-SECIR | An extension of the SECIR type implemented as input-output non-linear dynamical system. Joint fit of data on test positives, deaths, and ICU occupancy accounting for reporting delays. | ✓ |  |  | ✓ |  | ECDC | ✓ |
|  | MIMUW-StochSEIR | SEIR model with extensions: introduction of the undiagnosed compartment; testing limits influencing number of diagnosed cases; stochastic perturbations of time-dependent contact rate. |  |  |  |  | ✓ | JHU | ✓ |
|  | UCLA-SuEIR<sup>2</sup> | A variant of the SEIR model considering both untested and unreported cases (Zou et al., 2020). The model considers reopening and assumes the susceptible population will increase after the reopen. | ✓ | ✓ |  | ✓ |  | JHU |  |
|  | USC-SIkJalpha<sup>3</sup> | Reduces a heterogeneous rate model into multiple simple linear regression problems. True susceptible population is identified based on reported cases, whenever possible. (Srivastava et al., 2020). |  |  |  | ✓ | ✓ | JHU |  |
| Growth rate/<br>renewal eq. | epiforecasts-EpiNow2 | An exponential growth model that uses a time-varying $R_t$ trajectory to forecast latent infections, then convolves these using known delays to observations. (Abbott et al., 2020). Beyond the forecast horizon $R_t$ is assumed to be static. | | | | ✓ | ✓ | ECDC | ✓ |
|  | Geneva-DetGrowth<sup>4</sup> | Robust seasonal trend decomposition for smoothing of daily observations with further linear or multiplicative extrapolation. |  |  |  | ✓ | ✓ | ECDC |  |
|  | ITWW-county_repro | Forecasts of county level incidence based on regional reproduction numbers estimated via small area estimation. |  |  | ✓ | ✓ | ✓ | ECDC | ✓ |
|  | LANL-GrowthRate<sup>5</sup> | Dynamic SI model for cases with growth rate parameter updated at each model run (via regression model with day-of-week effect). The deaths forecast is a fraction of the cases forecasts (fraction learned via regression and updated at each run). |  |  |  | ✓ | ✓ | JHU | ✓ |
| Human judgement | epiforecasts-EpiExpert | A mean ensemble of predictions from experts and non-experts. Predictions are made through a web app<sup>6</sup> by choosing a distribution and specifying the median and width of that predictive distribution. | (✓) | (✓) | (✓) | ✓ | ✓ | ECDC | ✓ |
| Forecast ensemble | Imperial-ensemble2<sup>7</sup> | Unweighted average of four forecasts for death counts (see reference in footnote). |  |  |  | ✓ | ✓ | ECDC | ✓ |

Teams marked with footnotes run their own dashboards: <sup>1</sup>https://www.covidanalytics.io, <sup>2</sup>https://covid19.uclaml.org, <sup>3</sup>https://scc-usc.github.io/ReCOVER-COVID-19, <sup>4</sup>https://renkulab.shinyapps.io/COVID-19-Epidemic-Forecasting, <sup>5</sup>https://covid-19.bsvgateway.org, <sup>6</sup>https://cmmid-lshtm.shinyapps.io/crowd-forecast, <sup>7</sup>https://mrc-ide.github.io/covid19-short-term-forecasts

**Table 2:**
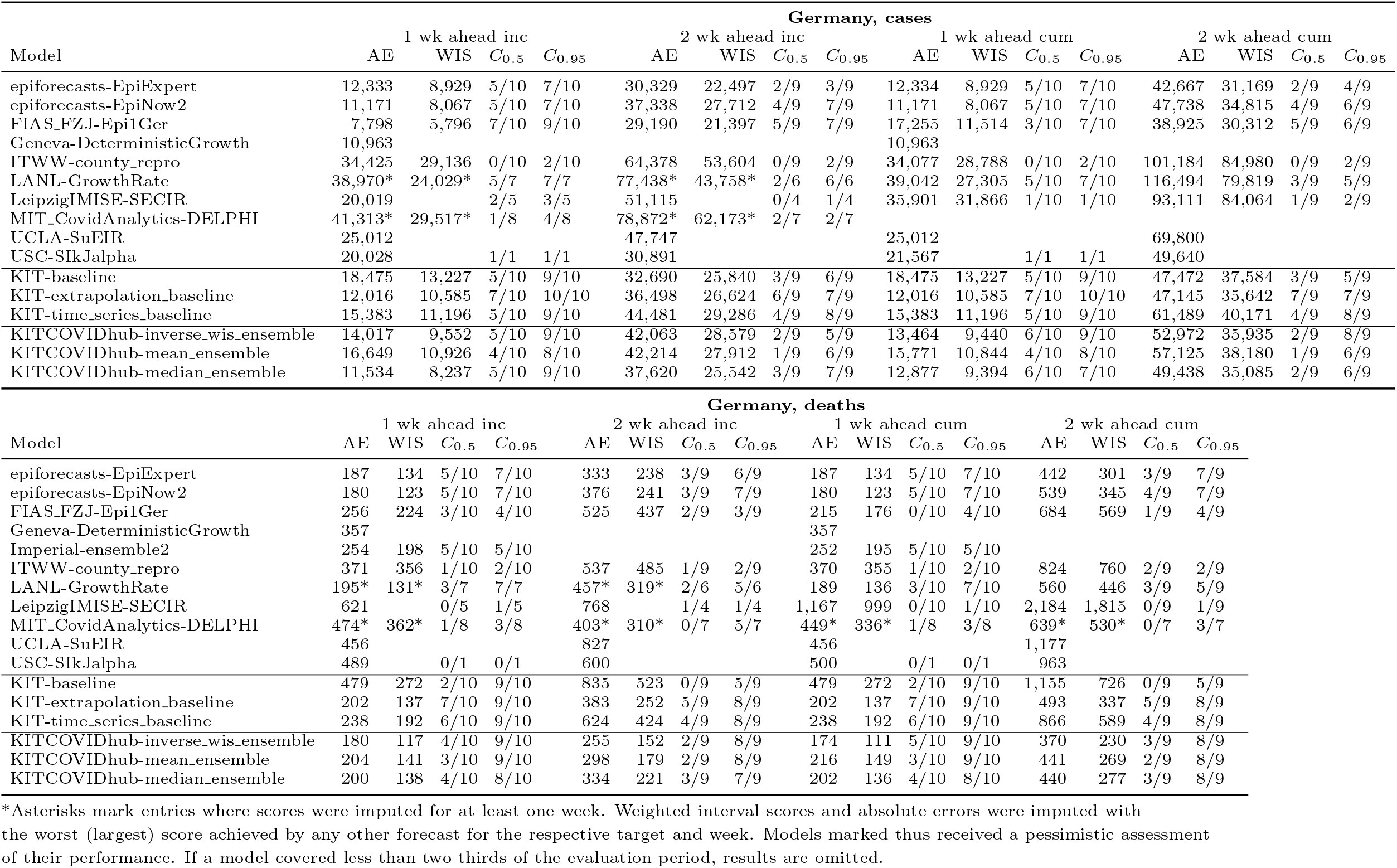
Detailed summary of forecast evaluation for Germany (based on ECDC data). *C*_0.5_ and *C*_0.95_ denote coverage rates of the 50% and 90% prediction intervals; AE and WIS stand for the mean absolute error and mean weighted interval score.

**Table 3:**
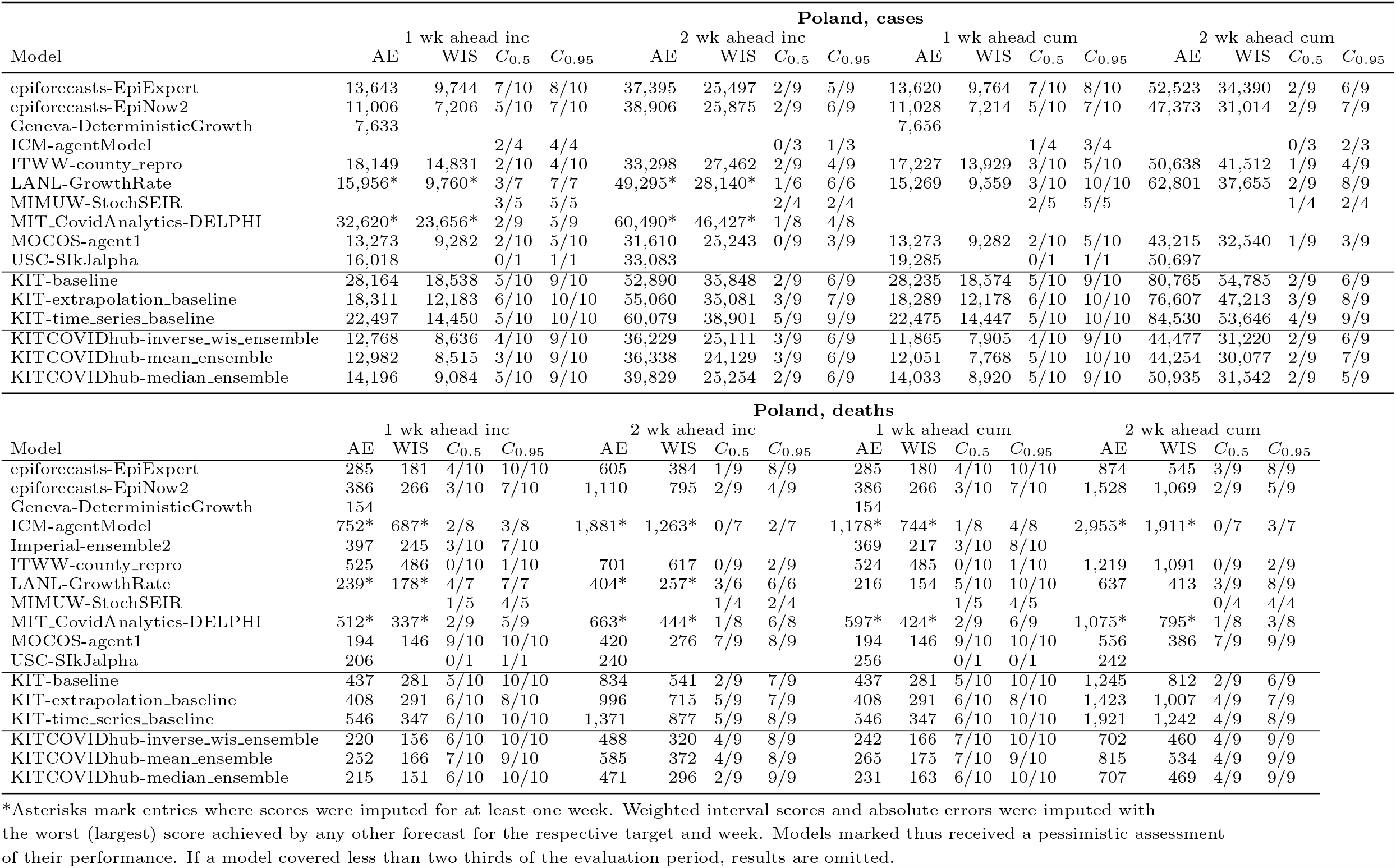
Detailed summary of forecast evaluation for Poland (based on ECDC data). *C*_0.5_ and *C*_0.95_ denote coverage rates of the 50% and 90% prediction intervals; AE and WIS stand for the mean absolute error and mean weighted interval score.

**Table 4:**
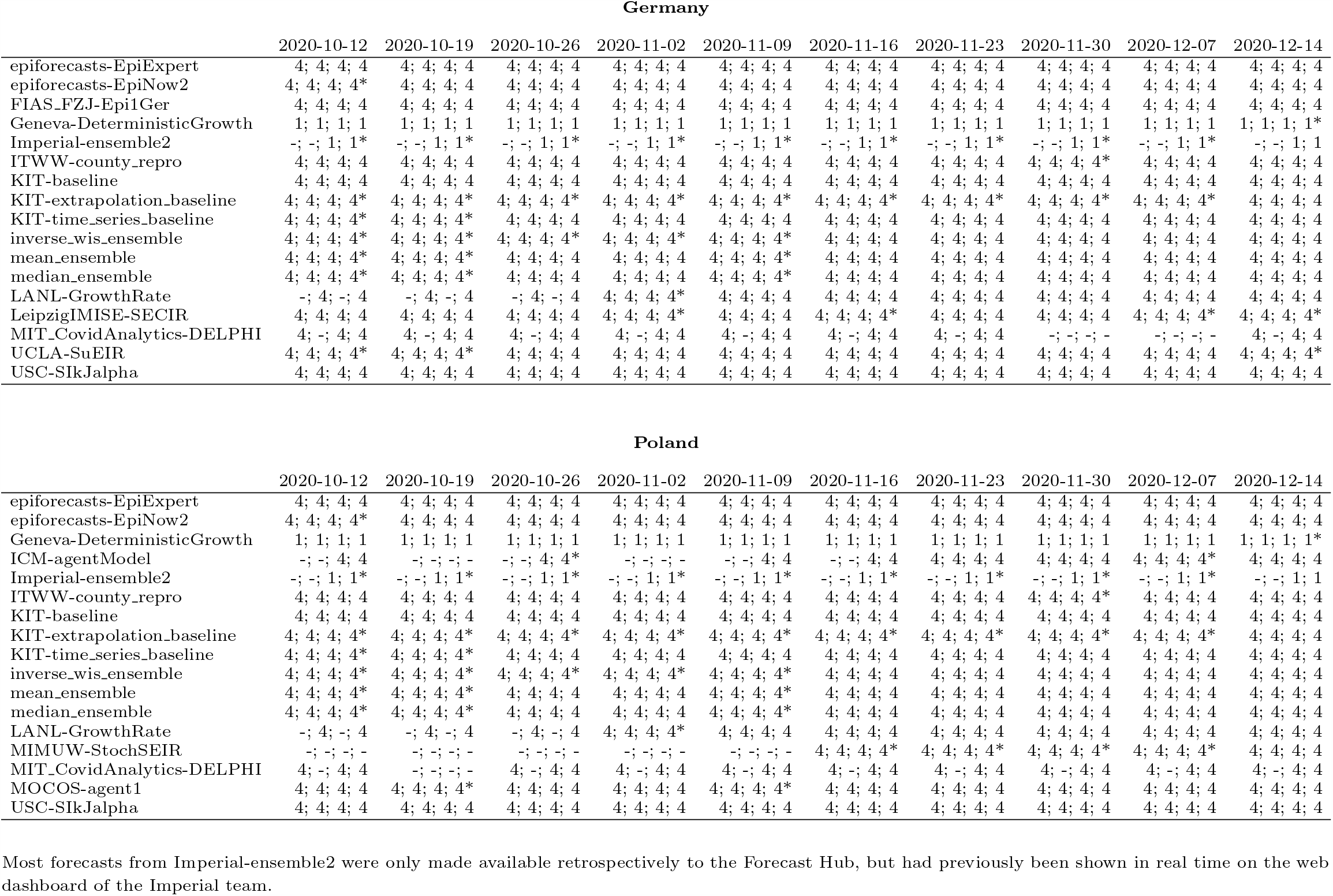
Availability of forecasts by model, target and forecast horizon.

**Table 5:**
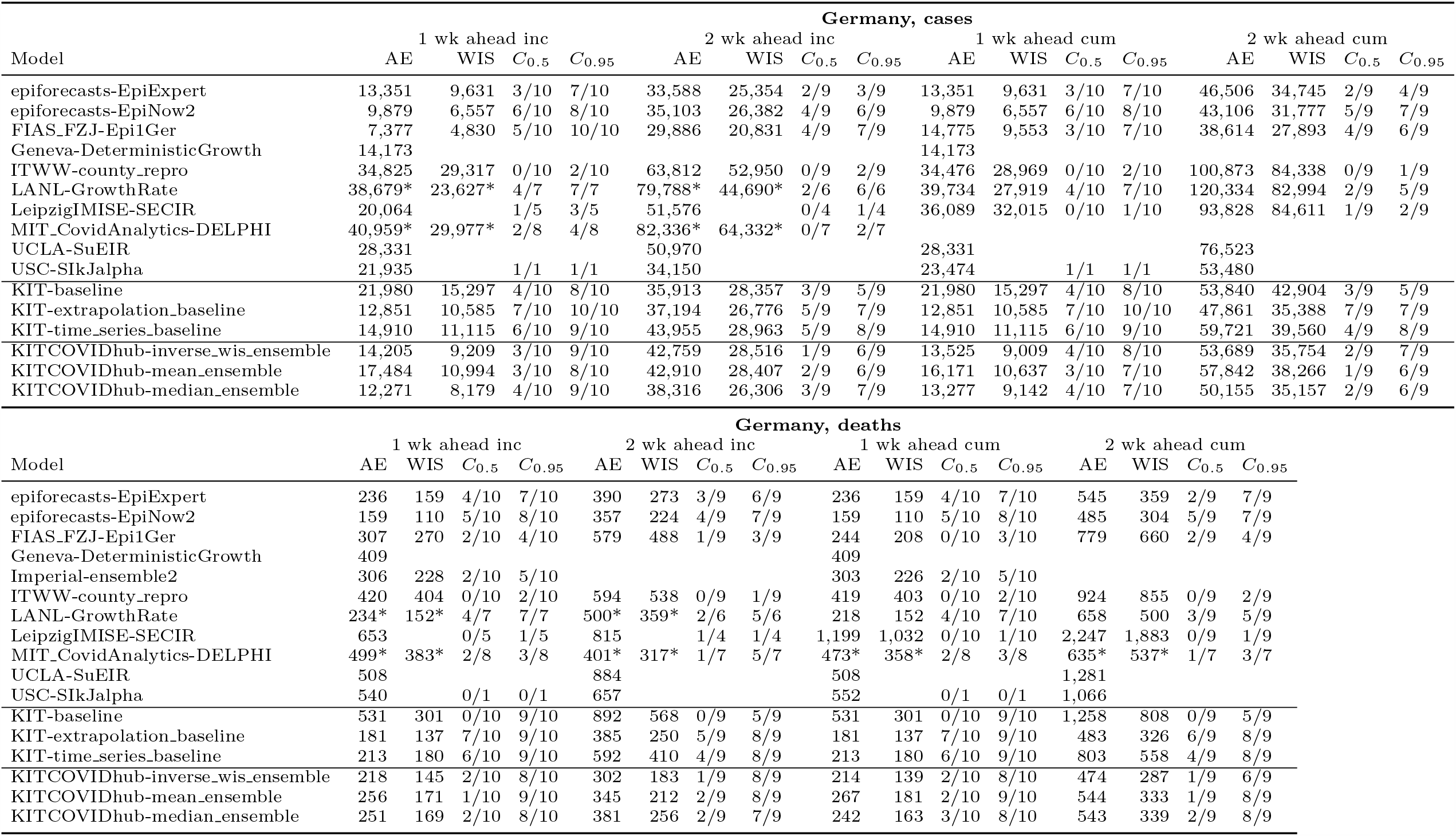
Detailed summary of forecast evaluation for Germany (based on JHU data)

**Table 6:**
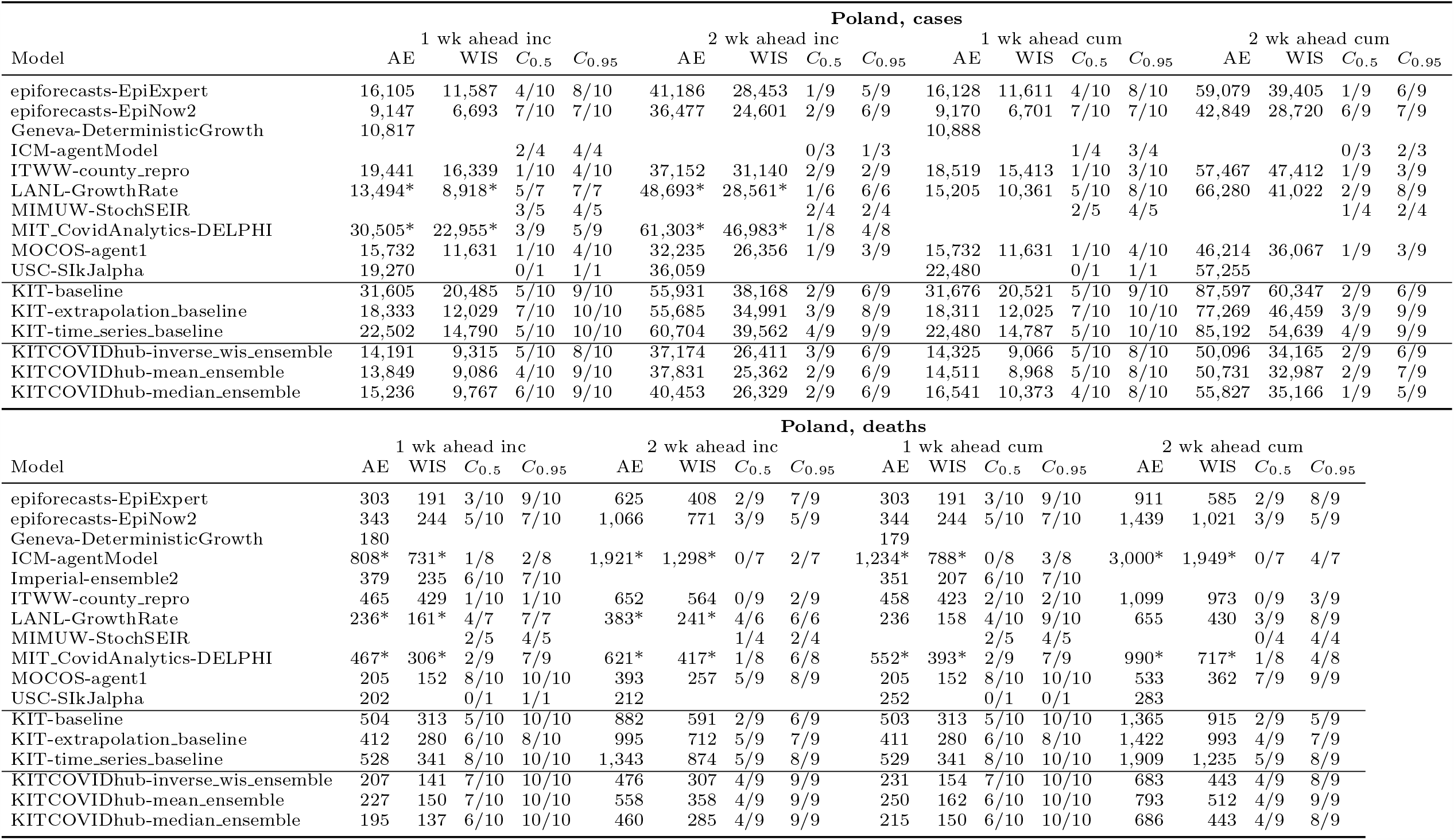
Detailed summary of forecast evaluation for Poland (based on JHU data)

**Table 7:**
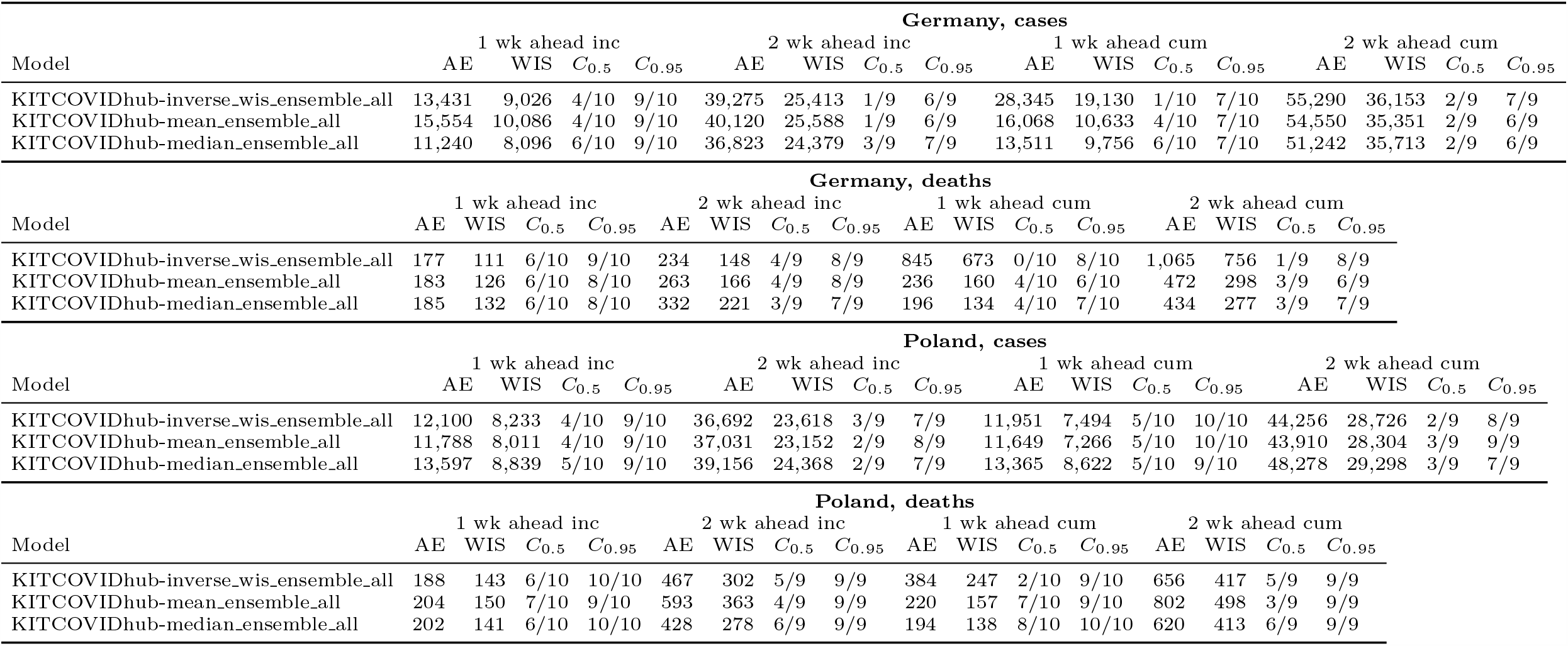
Summary of forecast evaluation for ensembles without plausibility checks of members (based on ECDC data)

**Table 8:**
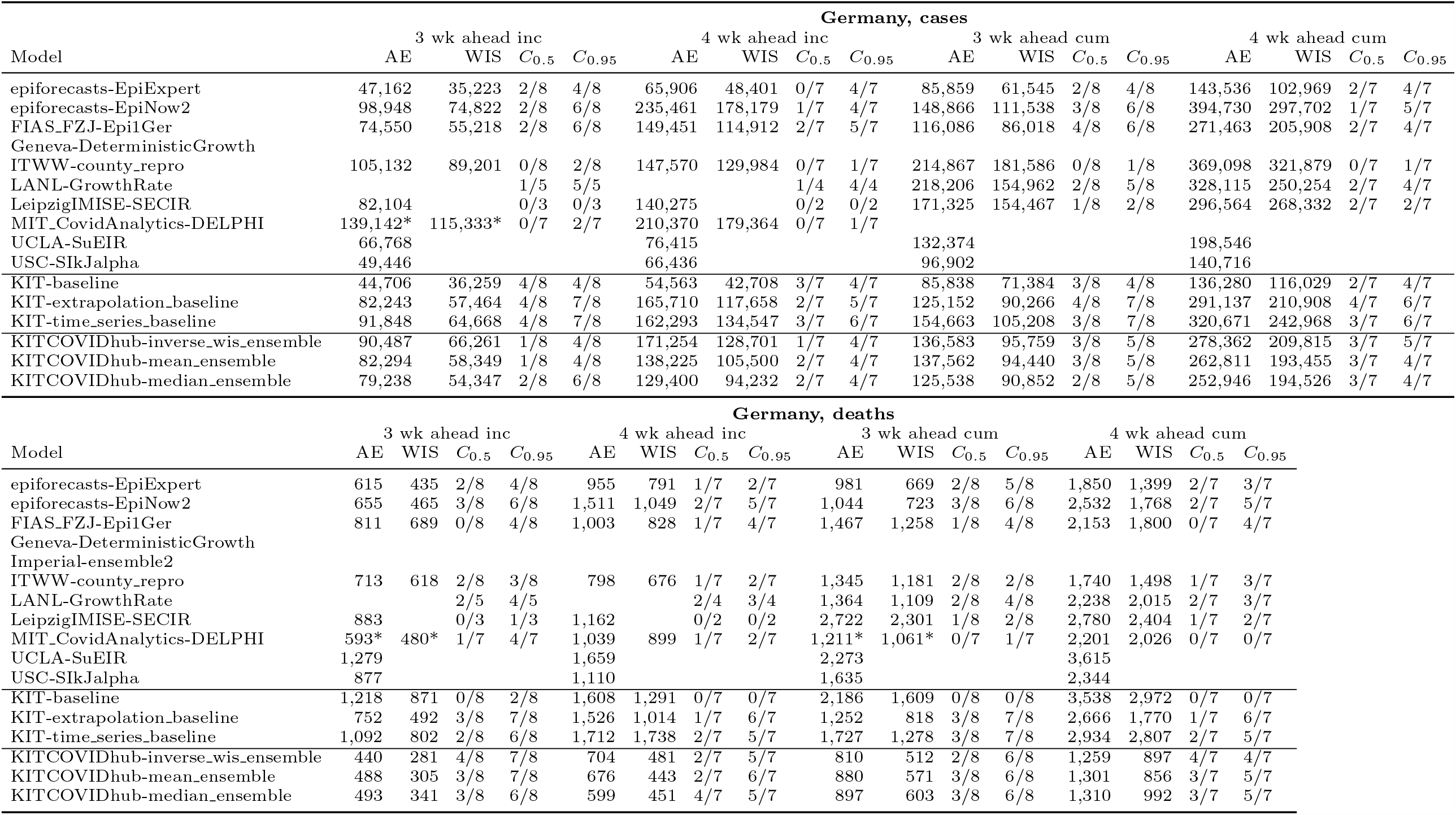
Detailed summary of forecast evaluation for Germany, 3 and 4 weeks ahead (based on ECDC data)

**Table 9:**
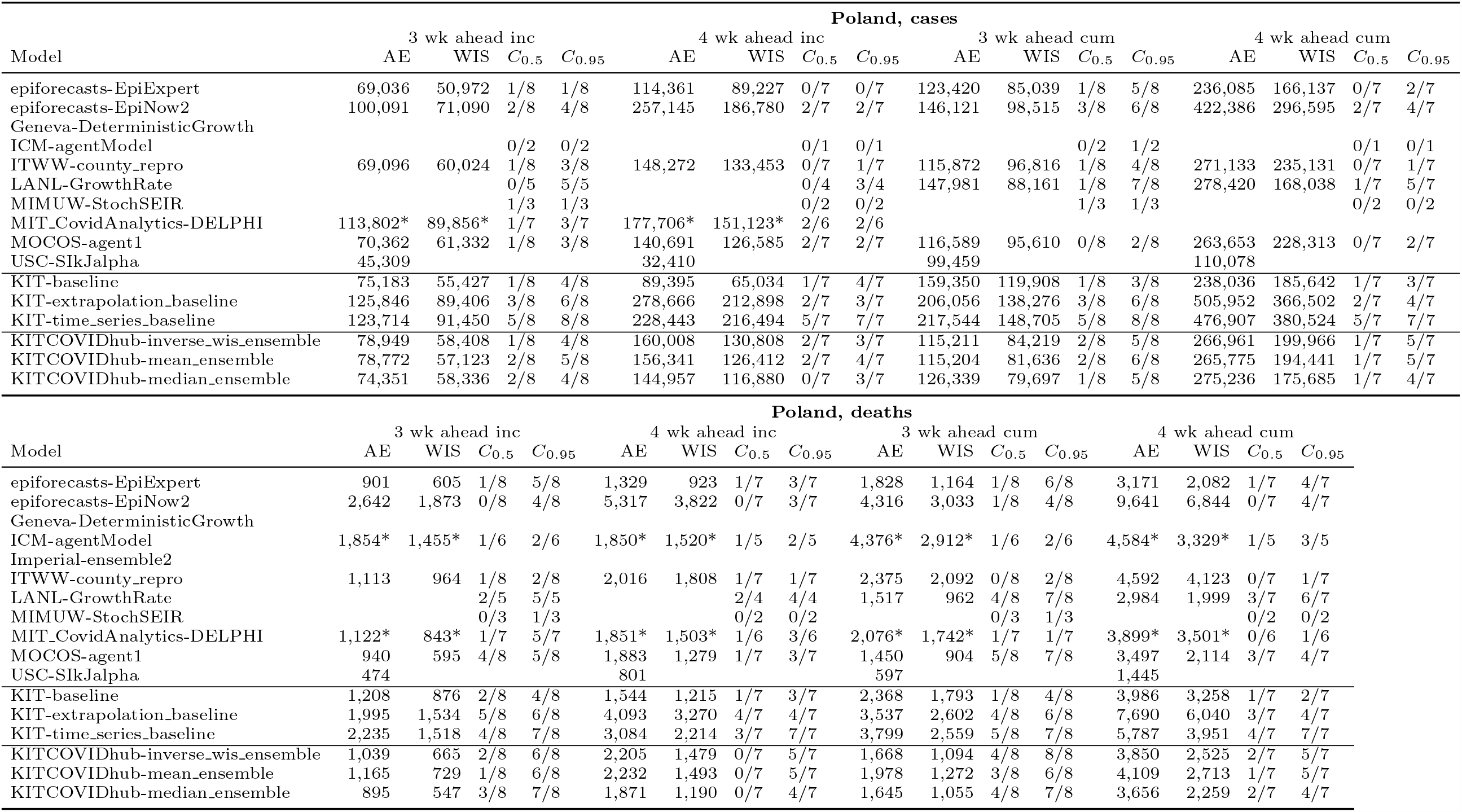
Detailed summary of forecast evaluation for Poland, 3 and 4 weeks ahead (based on ECDC data)

### 3.3 Ensemble forecasts

Evidence from past forecasting efforts on various diseases (e.g., Yamana et al. 2016; Viboud et al. 2018; Reich et al. 2019b) and recent research in the context of COVID-19 (Brooks et al., 2020; Funk et al., 2020) suggests that ensemble forecasts combining several independent forecasts can lead to improved and more stable performance. We therefore assess the performance of three different forecast aggregation approaches:

`KITCOVIDhub-median_ensemble` The *α*-quantile of the ensemble forecast for a given quantity is given by the median of the respective *α*-quantiles of the member forecasts. The associated point forecast is the quantile at level *α* = 0.50 of the ensemble forecast (same for other ensemble approaches).

`KITCOVIDhub-mean_ensemble` The *α*-quantile of the ensemble forecast for a given quantity is given by the mean of the respective *α*-quantiles of the member forecasts.

`KITCOVIDhub-inverse_wis_ensemble` The *α*-quantile of the ensemble forecast is a weighted average of the *α*-quantiles of the member forecasts. The weights are chosen inversely to the mean WIS value obtained by the member models over six recently evaluated forecasts (last three one-week-ahead, last two two-week-ahead, last three-week-ahead). This is done separately for incident and cumulative forecasts. The inverse-WIS ensemble is a pragmatic strategy to base weights on past performance which is feasible with a limited amount of historical forecast/observation pairs (see Zamo et al. 2020 for a similar approach).

Only models providing complete probabilistic forecasts with 23 quantiles for all four forecast horizons were included into the ensemble for a given target. It was not required that forecasts be submitted for both cumulative and incident targets, so that ensembles for incident and cumulative cases were not necessarily based on exactly the same set of models. The Forecast Hub Team reserved the right to screen and exclude member models in case of implausibilities. Decisions on inclusion were taken simultaneously for all three ensemble versions and were documented in the Forecast Hub platform. The main reasons for the exclusion of forecasts from the ensemble were forecasts in an implausible order of magnitude or forecasts with vanishingly small or excessive uncertainty. As it showed comparable performance to submitted forecasts, the `KIT-time_series_baseline` model was included in the ensemble forecasts in most weeks.

Preliminary results from the US COVID-19 Forecast Hub indicate better forecast performance of the median compared to the mean ensemble (Taylor and Taylor, 2020), and the median ensemble has served as the operational ensemble since 28 July 2020. Up to date, trained ensembles yield only limited, if any, benefits (Brooks et al., 2020). We therefore prespecified the median ensemble as our main ensemble approach. Note that in the context of influenza forecasting (Reich et al., 2019b), ensembles have been constructed by combining probability densities rather than quantiles. These approaches have somewhat different behaviour, but no general statement can be made which one yields better performance (Lichtendahl et al., 2013). As in our setting member forecasts were reported in a quantile format we resort to quantile-based methods for aggregation.

## 4 Results

We start by discussing some general observations made during the evaluation period, shedding light on challenges and particularities of collaborative real-time forecasting during a pandemic. Subsequently, we provide a quantitative evaluation in terms of WIS, AE, and interval coverage. Visualizations of one- and two-week-ahead forecasts on the incidence scale are displayed in Figures 3 and 4, respectively, and will be discussed in the following subsections. Note that these figures are restricted to models submitted over (almost) the entire evaluation period and providing complete forecasts including 23 predictive quantiles. Forecasts from the remaining models are illustrated in Supplementary Section E. Forecasts at prediction horizons of three and four weeks are shown in Supplementary Section F.

### 4.1 Specific observations and challenges

A recurring theme during the evaluation period was pronounced variability between model forecasts. Figure 2 illustrates this aspect for point forecasts of incident cases in Germany, but it also holds for Poland and death forecasts. The left panel shows the spread of forecasts issued on 19 October 2020 and valid one to four weeks ahead. The models present very different outlooks, ranging from a return to the lower incidence of previous weeks to exponential growth. The graph also illustrates the difficulty of forecasting cases more than two weeks ahead. Several models had correctly picked up the upwards trend, but presumably a combination of the new testing regime and the semi-lockdown (marked as (a) and (b)) led to a flattening of the curve. The right panel shows forecasts from 9 November 2020, immediately following the aforementioned events. Again, the forecasts are quite heterogeneous. The week ending on Saturday 7 November had seen a slower increase in reported cases than anticipated by almost all models (see Figure 3), but there was general uncertainty about the role of saturating testing capacities and evolving testing strategies. Indeed, on 18 November it was argued in a situation report from Robert Koch Institute (RKI) that comparability of data from calendar week 46 (9–15 November) to previous weeks was limited (Robert Koch Institute, 2020). This illustrates that confirmed cases can be a moving target, and that different modelling decisions can lead to very different forecasts.

**Figure 2:**
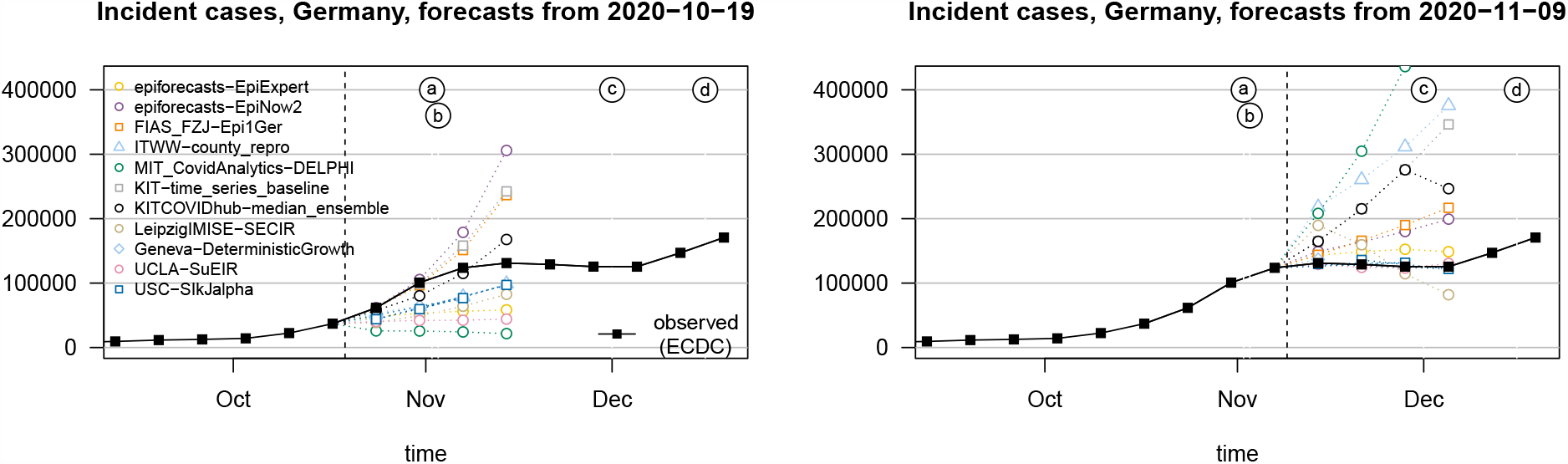
Illustration of heterogeneity between incident case forecasts in Germany. Left: Point forecasts issued by different models and the median ensemble on 19 October 2020. Right: Point forecasts issued on 9 November 2020. The dashed vertical line indicates the date at which forecasts were issued. Events marked by letters a – d are explained in Figure 1.

Far from all forecast models explicitly account for interventions and testing strategies (Table 1). Many forecasters instead prefer to let their models pick up trends from the data once they become apparent. This can lead to delayed adaptation to changes and explains why numerous models – including the ensemble – showed overshoot in the first half of November when cases started to plateau in Germany (visible from Figure 3 and even more pronounced in Figure 4). Interestingly, some models adapted more quickly to the flatter curve. This includes the human judgement approach `EpiExpert`, which, due to its reliance on human input and knowledge, can take information on interventions into account before they become apparent in epidemiological data, but interestingly also `Epi1Ger` and `EpiNow2` which do not account for interventions. In Poland, overshoot could be observed following the peak week in cases (ending on 15 November), with the one-week-ahead median ensemble only barely covering the next observed value. However, most models adapted quickly and were back on track in the following week.

**Figure 3:**
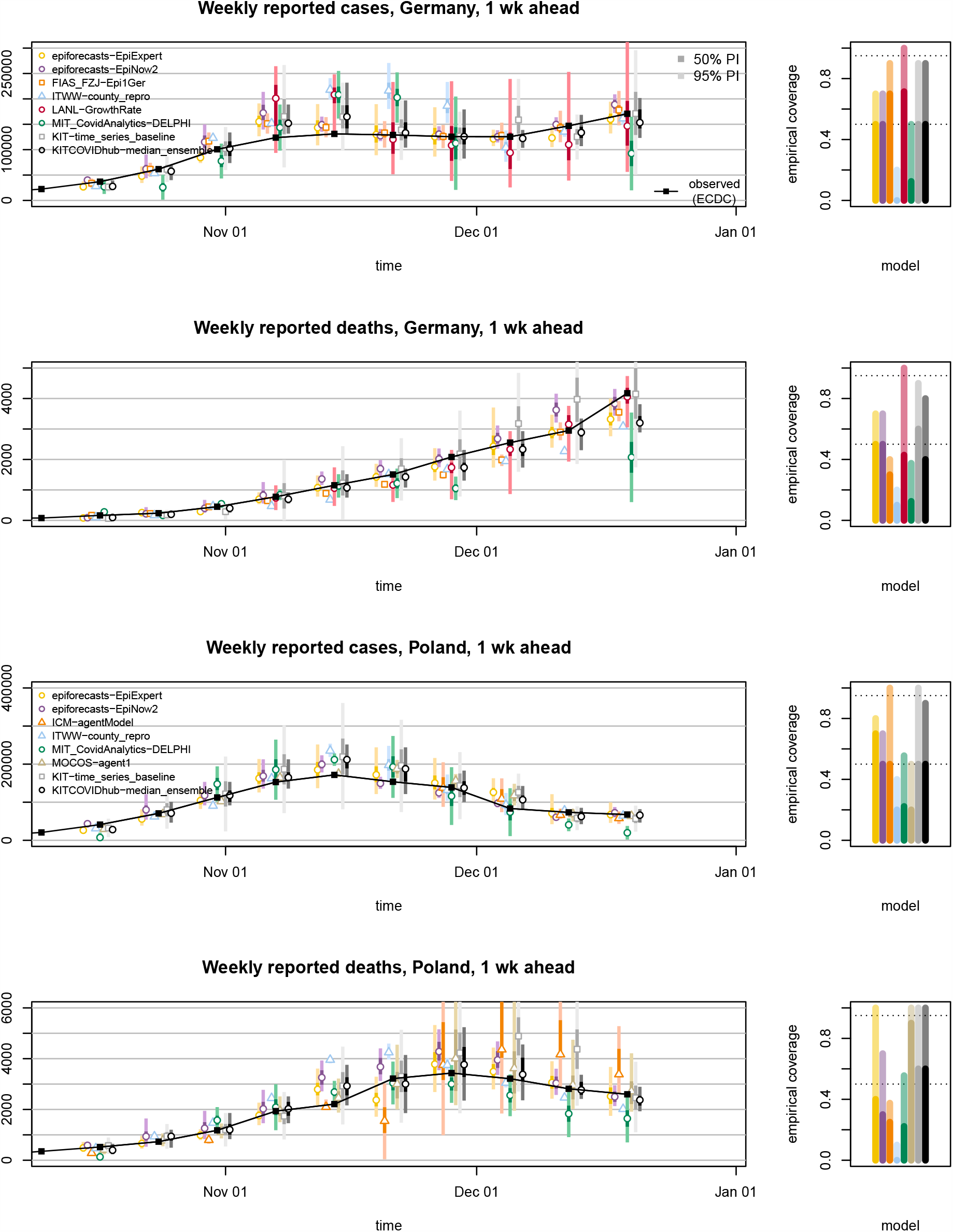
One-week-ahead forecasts of incident cases and deaths in Germany and Poland (left column). Displayed are predictive medians, 50% and 95% prediction intervals. Coverage plots (right column) show the empirical coverage of 95% (light) and 50% (dark) prediction intervals.

**Figure 4:**
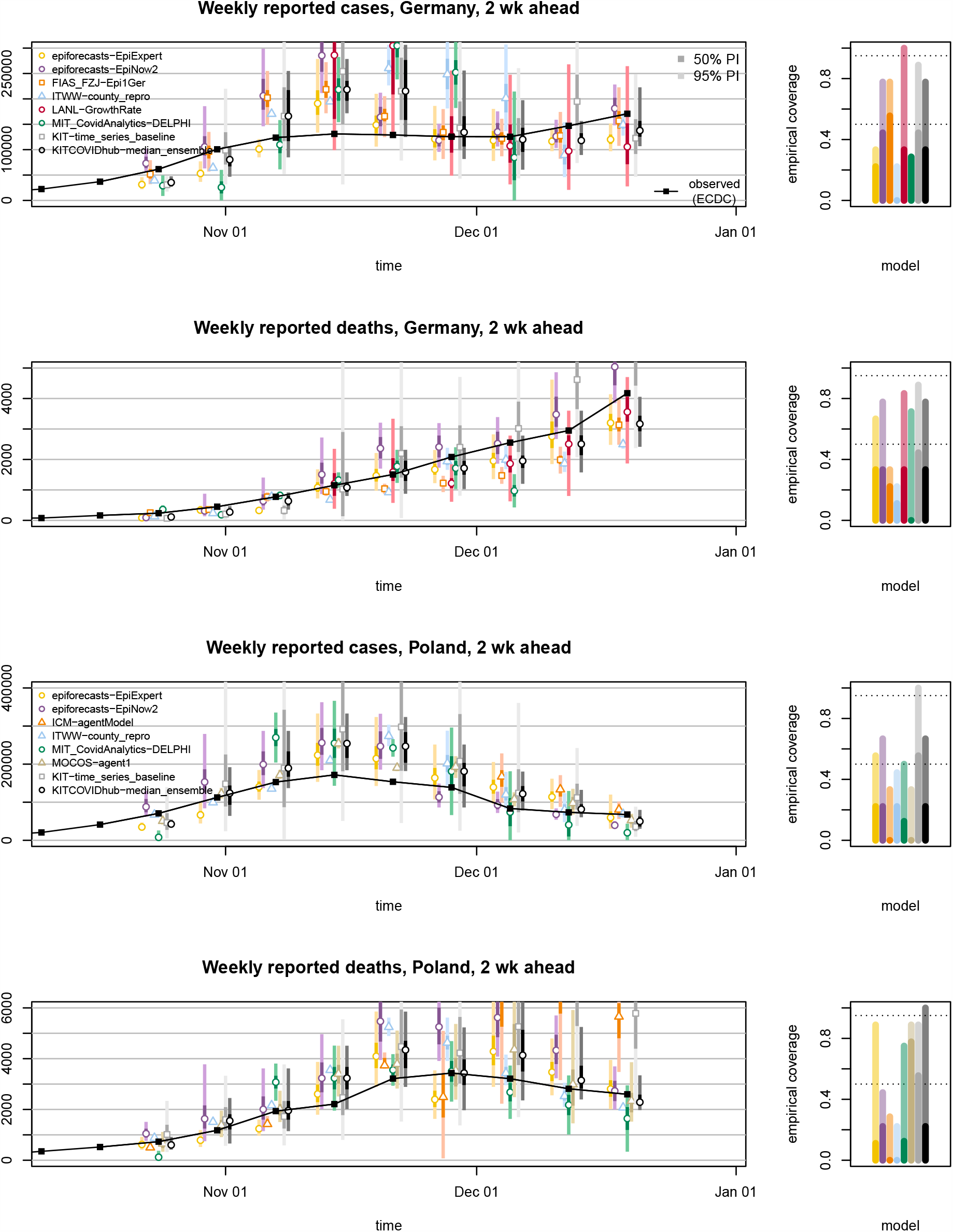
Two-week-ahead forecasts of incident cases and deaths in Germany and Poland (left column). Displayed are predictive medians, 50% and 95% prediction intervals. Coverage plots (right column) show the empirical coverage of 95% (light) and 50% (dark) prediction intervals.

A noteworthy difficulty for death forecasts in Germany was under-prediction in consecutive weeks in late November and December. In November, several models predicted that death numbers would stop increasing, likely as a consequence of the plateau in overall case numbers starting several weeks before. In the last week of our study (ending on 19 December) most models considerably under-estimated the increase in weekly deaths. A difficulty may have been that despite the overall plateau which was observed until early December, cases continued to increase in the oldest age groups, for which the mortality risk is highest (see Supplementary Figure 8). Models that do not take into account the age structure of cases – which includes most available models (Table 1) – may then have been led astray.

Forecasts are not only heterogeneous with respect to their point forecasts, but also the implied uncertainty. As can be seen from Figures 3 and 4, certain models issue very confident forecasts with narrow forecast intervals barely visible in the plot. Others – in particular the exponential smoothing time series model `KIT-time_series_baseline`, but also `LANL-GrowthRate` – show rather large uncertainty. For almost all forecast dates there are pairs of models with no or minimal overlap in 95% prediction intervals, another indicator of limited agreement between forecasts. As can be seen from the right column of Figures 3 and 4 as well as Tables 2 and 3, most contributed models were overconfident, i.e. their prediction intervals did not reach nominal coverage.

A major question in pandemic real-time forecasting is how closely surveillance data reflect the underlying dynamics. Like in Germany, testing criteria were repeatedly adapted in Poland. In early September they were tightened, requiring the simultaneous presence of four symptoms for the administration of a test. This was changed to less restrictive criteria in late October (presence of one characteristic symptom alone sufficient). These changes limit comparability of numbers across time. Very high test positivity rates in Poland suggest that there was substantial under-ascertainment, which is assumed to have aggravated over time. Comparisons between overall excess mortality and reported COVID deaths suggest that there is also relevant under-ascertainment of deaths, again likely changing over time (Afelt et al., 2020). These aspects make predictions challenging, and limitations of ground truth data sources are inherited by the forecasts which refer to them. A particularly striking example of this was the belated addition of 22,000 cases from previous weeks to the Polish record on 24 November 2020. We are aware that certain teams (namely, the Poland-based teams `MOCOS` and `MIMUW`) explicitly took this shift into account while others did not. This incident was not specifically accounted for in the evaluation as it was considered part of the general uncertainty affecting the prediction targets.

### 4.2 Findings for median, mean and inverse-WIS ensembles

Beyond comparing and evaluating short-term forecasts, we assessed the potential of forecast ensembles. Before providing a quantitative assessment in the following section, we present some general observations on the median, mean and inverse-WIS ensembles introduced in Section 3.3.

A key advantage of the median ensemble is that it is more robust to single extreme forecasts than the mean ensemble. As an example of the different behaviour in cases where one forecast differs considerably from the others we show forecasts of incident deaths in Poland from 30 November 2020 in Figure 5. The first panel shows the six member forecasts, the second the resulting median and mean ensembles. While the two ensemble forecasts are not drastically different and imply rather similar ranges, the predictive median of the latter is noticeably higher. The reason is that it is more strongly impacted by one model which predicted a resurge in deaths.

**Figure 5:**
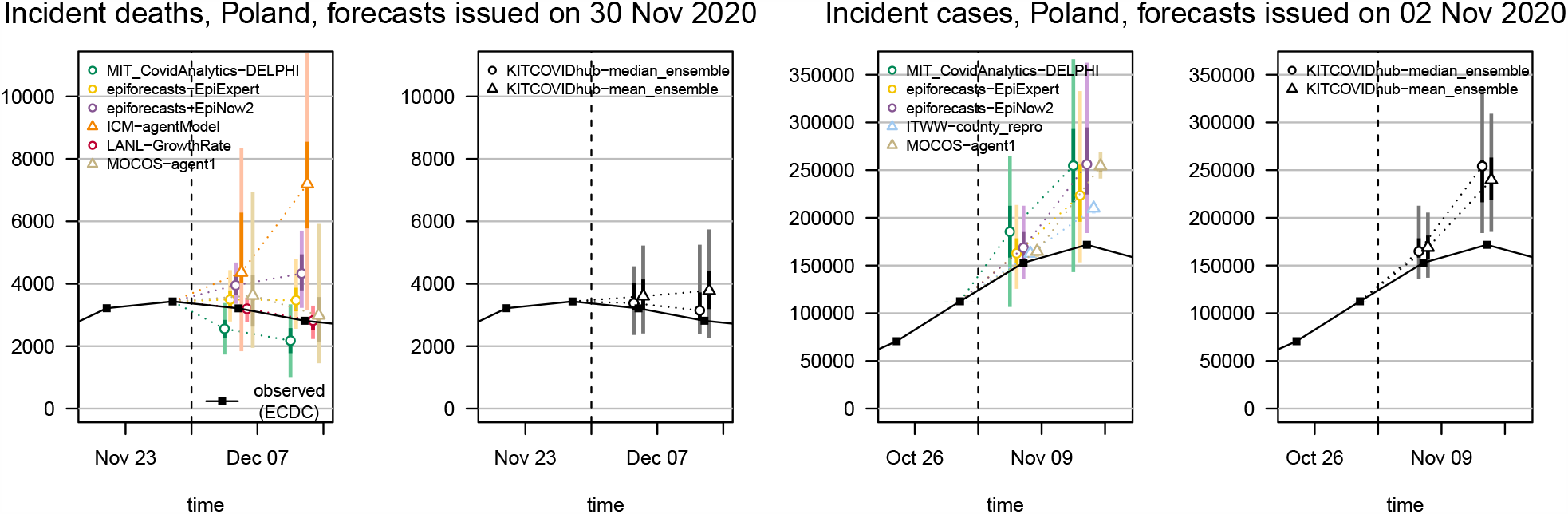
Examples of median and mean ensembles: One- and two-week-ahead forecasts of incident deaths in Poland issued on 30 November (first and second panel), and of incident cases in Poland issued on 2 November 2020 (third and fourth panel). The first and third panels show the member forecasts, the second and fourth panels the respective ensembles. Both predictive medians and 95% (light) and 50% (dark) prediction intervals are shown. The dashed vertical line indicates the date at which the forecasts were issued.

While the robustness of the median ensemble is often an advantage, we also encountered a downside of the approach. When member forecasts are rather heterogeneous, and there are low to medium numbers of members only, median ensemble forecasts are not always very well-shaped. One of the most pronounced examples we encountered is shown in the third and fourth panel of Figure 5. For the one-week-ahead forecast of incident cases in Poland from 2 November 2020, the predictive 25% quantile and median were almost identical. For the two-week-ahead median ensemble forecast, the 50% and 75% quantile were almost identical. Both distributions are thus rather oddly shaped and not a very plausible belief about the future, with a quarter of the probability mass concentrated in a very short interval. The mean ensemble, on the other hand, produces a more symmetric and thus more realistic representation of the associated uncertainty.

We now briefly address the inverse-WIS ensemble, which is a pragmatic approach to giving more weight to forecasts with good recent performance. Figure 6 shows the weights of the various member models for incident deaths in Germany and Poland. Note that in the first week, numerous models received the same weight as they were submitted for the first time and their scores for past weeks were all imputed with the same values (the worst scores achieved by any model in the respective week). While there are some models which on average receive larger weights than others, weights change considerably over time. Some models are not included in the ensemble for certain weeks, either because of delayed or missing submissions or due to concerns about their plausibility (Section 3.3). The pronounced changes in weights indicate that relative performance fluctuates over time, making it challenging to improve performance of ensemble forecasts by taking past results into account. A possible reason is that models get updated continuously by their maintainers, including major revisions of methodology. Indeed, the overall results shown in Tables 2 and 3 do not indicate any systematic benefits from inverse-WIS weighting.

**Figure 6:**
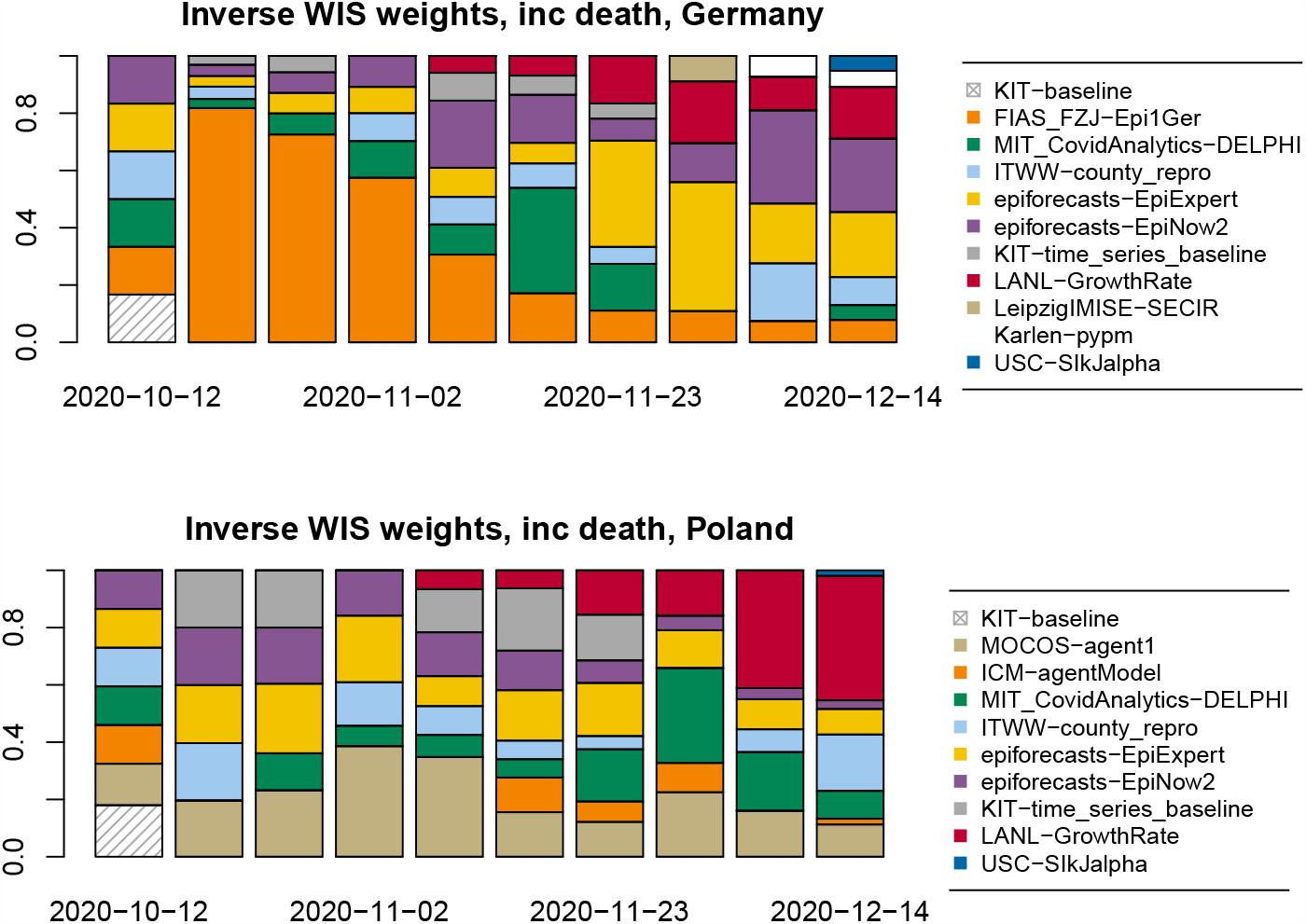
Inverse-WIS weights for forecasts of incident deaths in Germany (top) and Poland (bottom)

### 4.3 Formal forecast evaluation

Forecasts were evaluated using the mean weighted interval score (WIS), mean absolute error (AE) and interval coverage rates. Tables 2 and 3 provide a detailed overview of results by country, target and forecast horizon. We repeated all evaluations using JHU data as ground truth (shown in the Supplement), and the overall results seem robust to this choice. We also provide the same tables for three- and four-week-ahead forecasts in Supplementary Section F, though in view of the discussion in Section 2.2 their usability is limited.

Figure 7 depicts the mean WIS achieved by the different models on the incidence scale. For models providing only point forecasts, the mean AE is shown, which as mentioned in Section 2.3 can be compared to mean WIS values. For deaths, the ensemble forecasts and several submitted models outperform the baseline up to three or even four weeks ahead. As argued before, deaths are a more strongly lagged indicator, which favours predictability at somewhat longer horizons. Another aspect may be that at least in Germany, death numbers have been following a rather uniform upward trend over the study period, making it relatively easy to beat the baseline model. For cases, which are a more immediate measure, almost none of the compared approaches meaningfully outperformed the naïve baseline beyond a horizon of one or two weeks. Especially in Germany this result is largely due to the pronounced overshoot of forecasts in early November as discussed in Section 4.1. The `KIT-baseline` forecast by definition always predicts a plateau, which is what was observed in Germany for roughly half of the evaluation period. Good performance of the baseline is thus less surprising. Nonetheless, these results underscore that in periods of evolving intervention measures meaningful case forecasts are limited to a rather short time window. In this context we also note that the additional baselines `KIT-extrapolation_baseline` and `KIT-time_series_baseline` do not systematically outperform the naïve baseline and for most targets are neither among the best nor the worst performing approaches.

**Figure 7:**
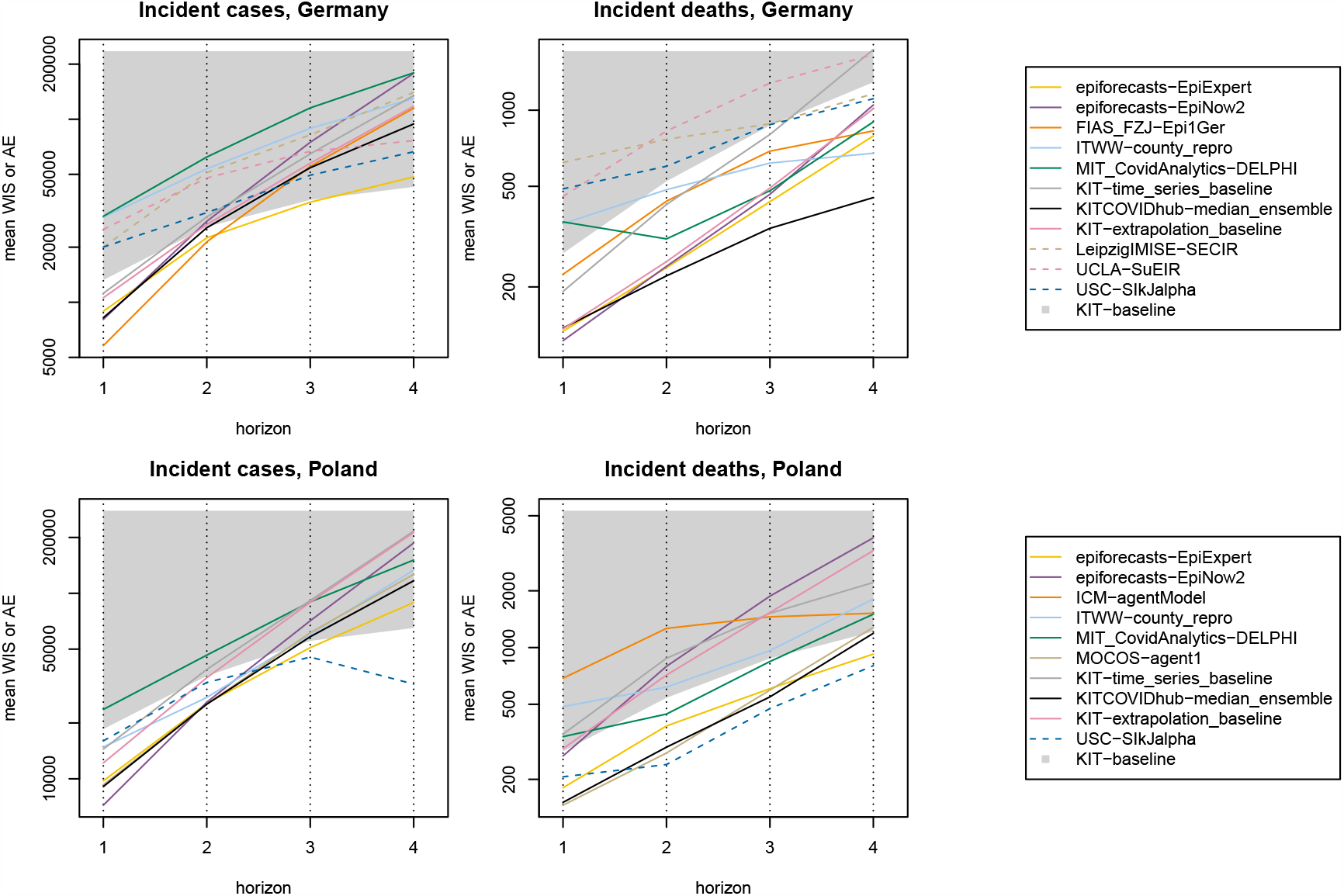
Mean WIS by forecast target and prediction horizon for submitted models and the preregistered median ensemble. For models providing only point forecasts, the mean AE is shown. The lower boundary of the grey area represents the baseline model `KIT-baseline`. Lines crossing the grey area thus indicate that a model fails to outperform the baseline. The numbers underlying this figure can also be found in Tables 2 and 3.

**Figure 8:**
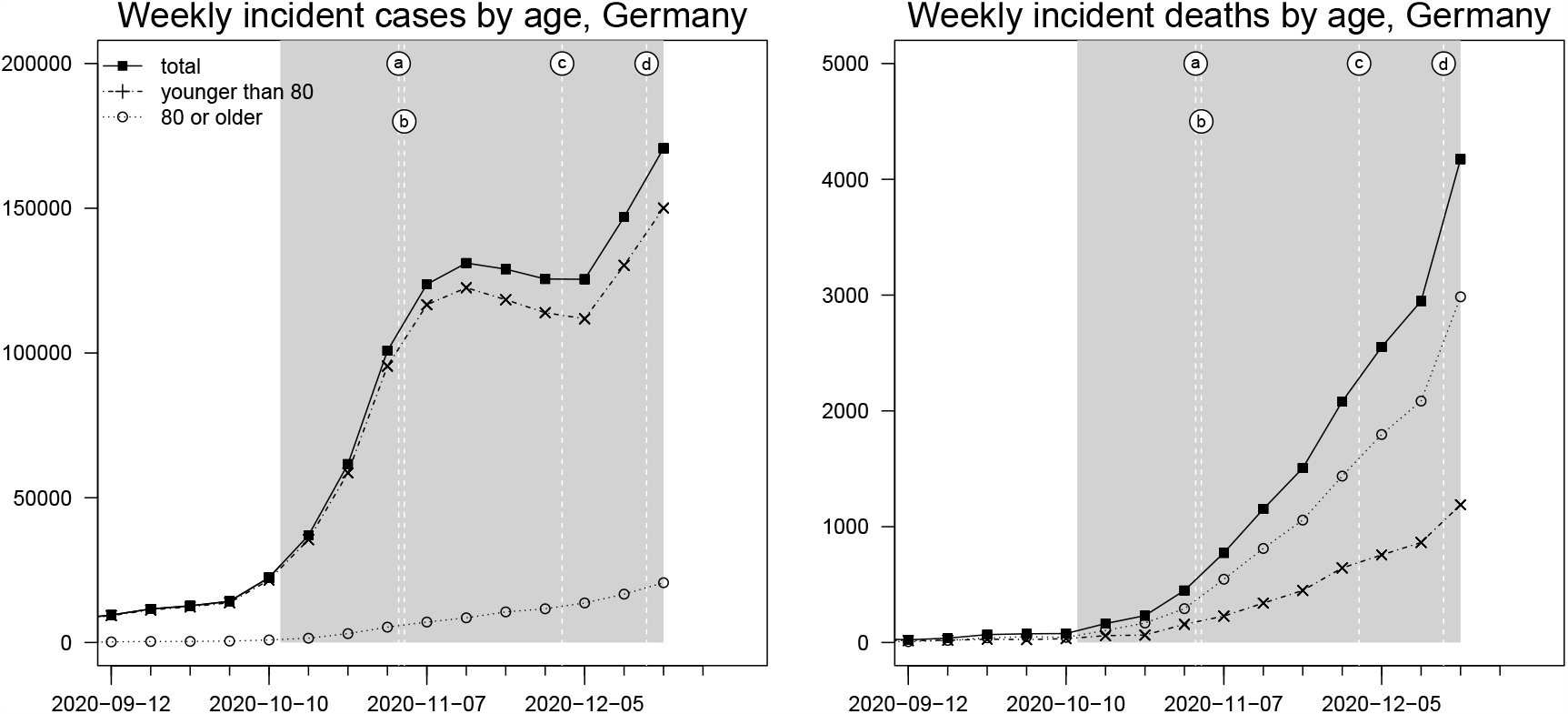
Weekly incident COVID-19 cases and deaths in Germany, pooled and stratified by age below and above 80 years. Events marked by letters a – d are explained in Figure 1.

**Figure 9:**
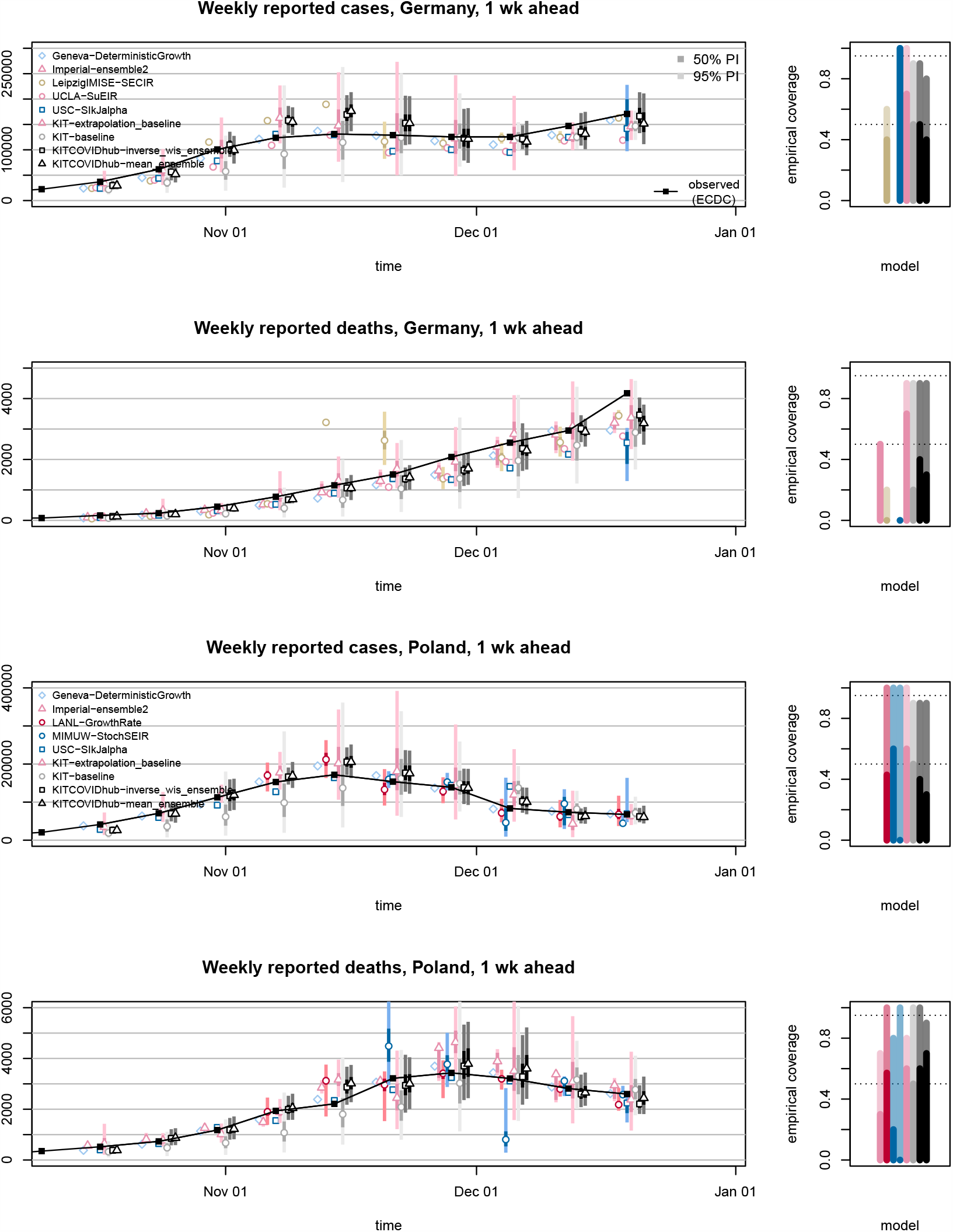
One-week-ahead forecasts of incident cases and deaths in Germany and Poland (left). Displayed are predictive medians, 50% and 95% prediction intervals for models not shown in Figure 3. Coverage plots (right) show the empirical coverage of 95% (light) and 50% (dark) prediction intervals.

**Figure 10:**
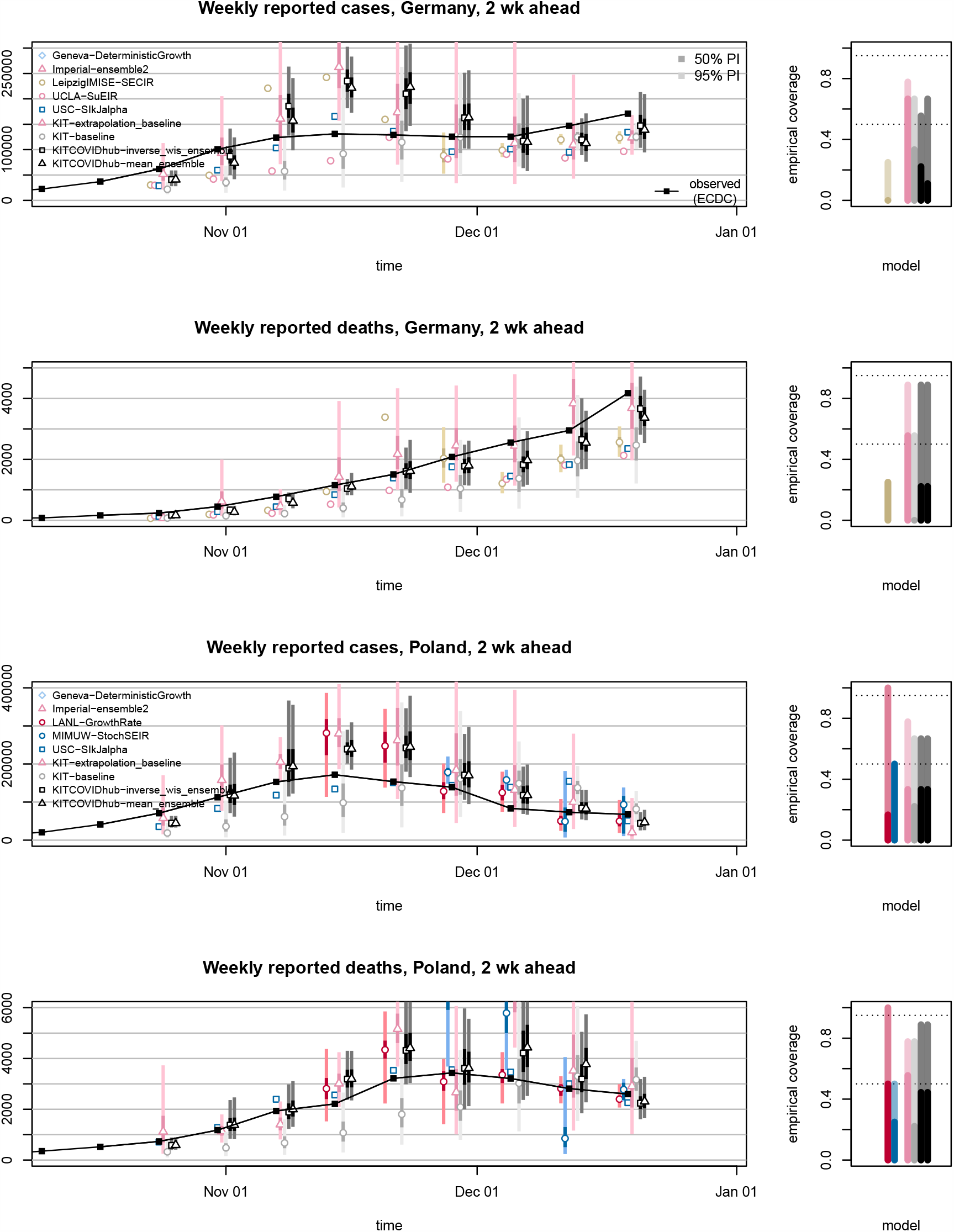
Two-week-ahead forecasts of incident cases and deaths in Germany and Poland (left). Displayed are predictive medians, 50% and 95% prediction intervals for models not shown in Figure 4. Coverage plots (right) show the empirical coverage of 95% (light) and 50% (dark) prediction intervals.

**Figure 11:**
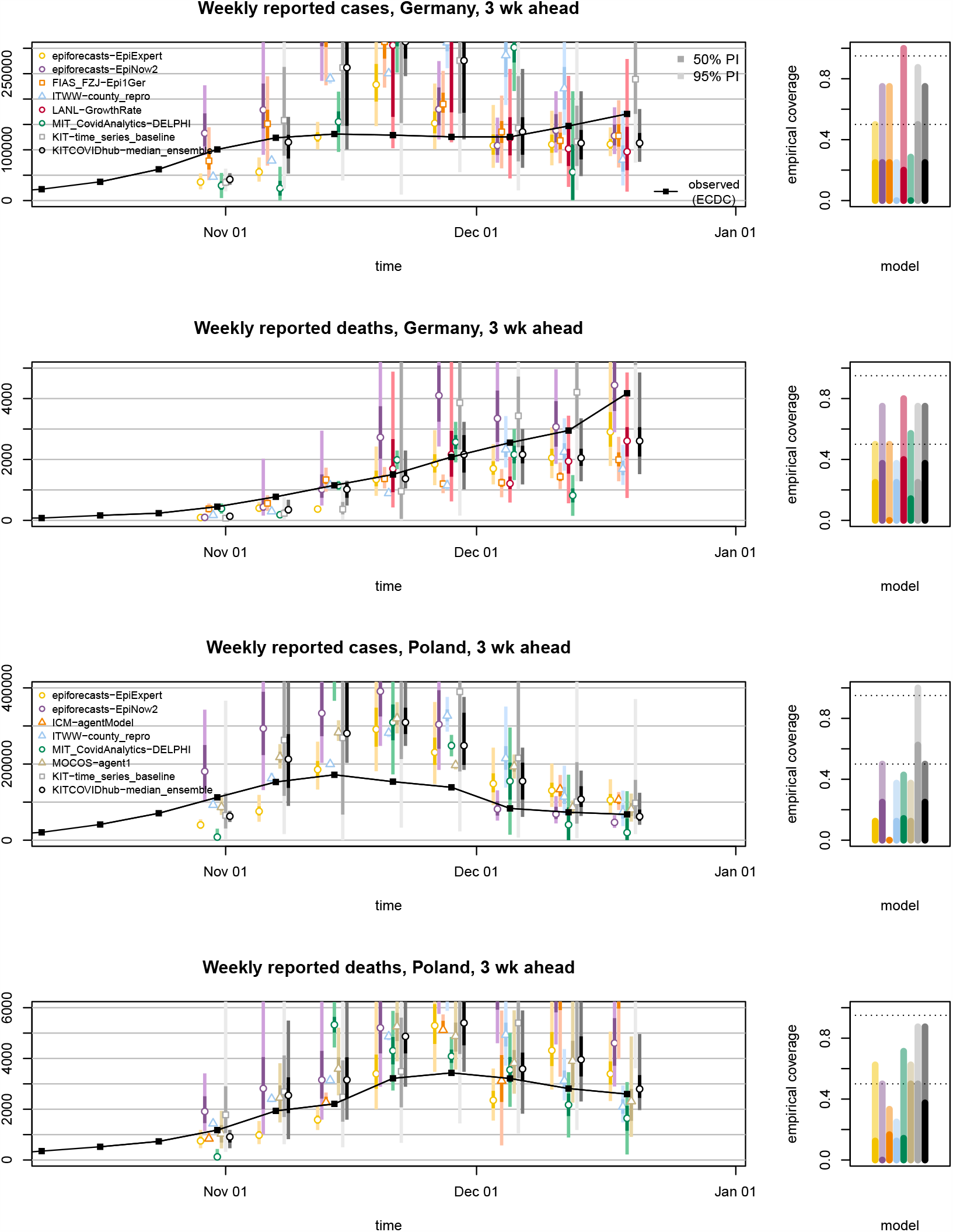
Three-week-ahead forecasts of incident cases and deaths in Germany and Poland (left). Displayed are predictive medians, 50% and 95% prediction intervals. Coverage plots (right) show the empirical coverage of 95% (light) and 50% (dark) prediction intervals.

**Figure 12:**
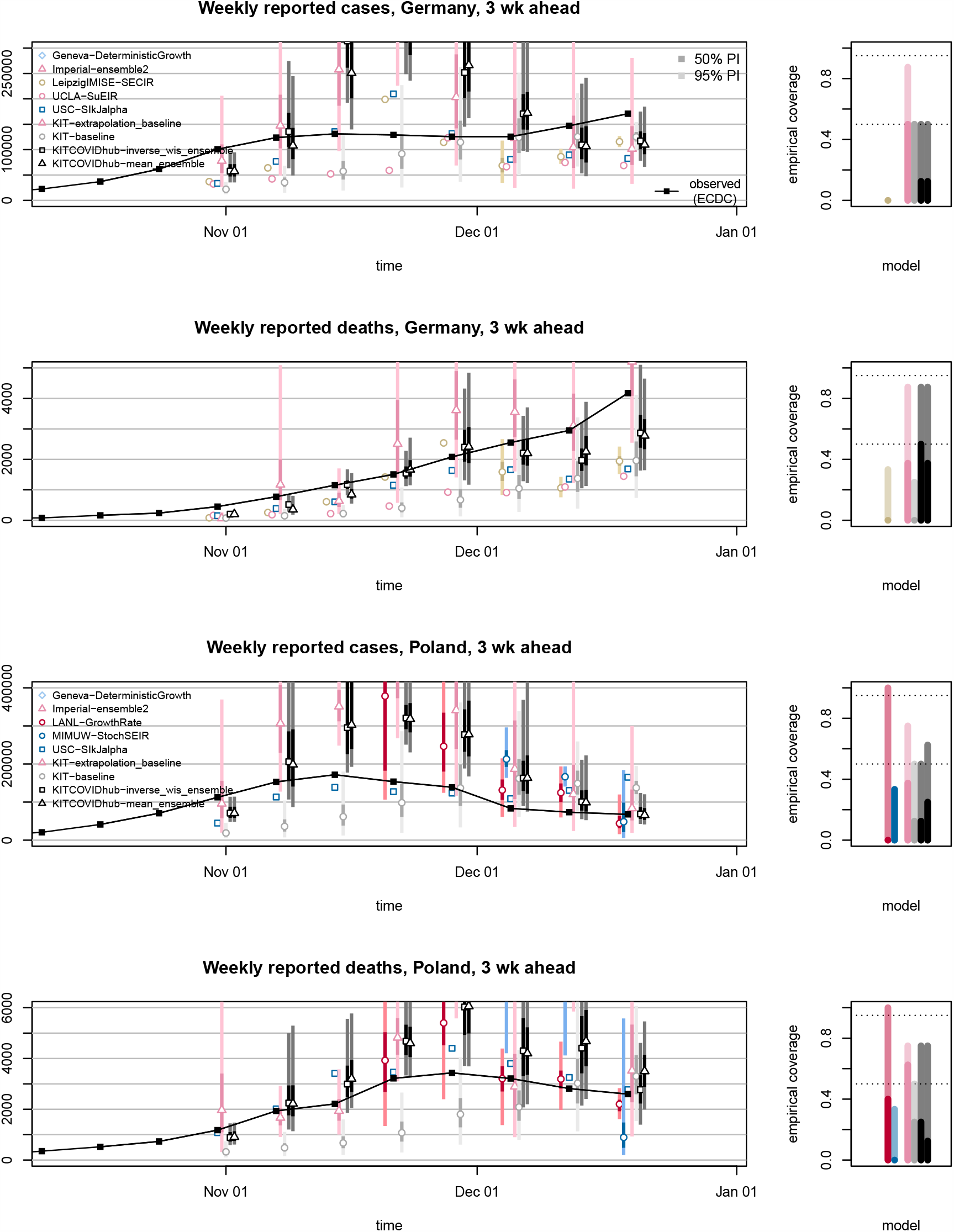
Three-week-ahead forecasts of incident cases and deaths in Germany and Poland (left). Displayed are predictive medians, 50% and 95% prediction intervals for models not shown in Figure 11. Coverage plots (right) show the empirical coverage of 95% (light) and 50% (dark) prediction intervals.

**Figure 13:**
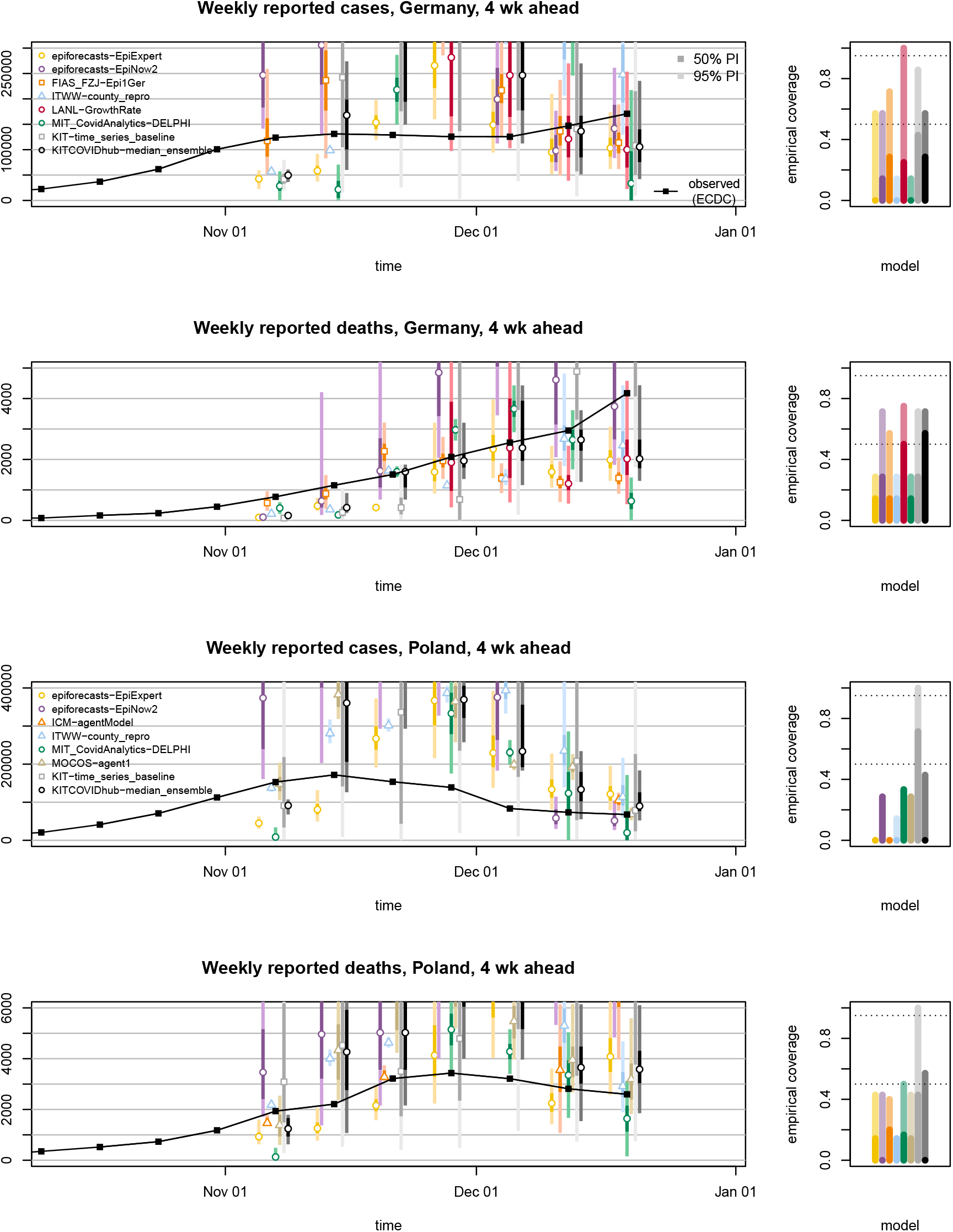
Four-week-ahead forecasts of incident cases and deaths in Germany and Poland (left). Displayed are predictive medians, 50% and 95% prediction intervals. Coverage plots (right) show the empirical coverage of 95% (light) and 50% (dark) prediction intervals.

**Figure 14:**
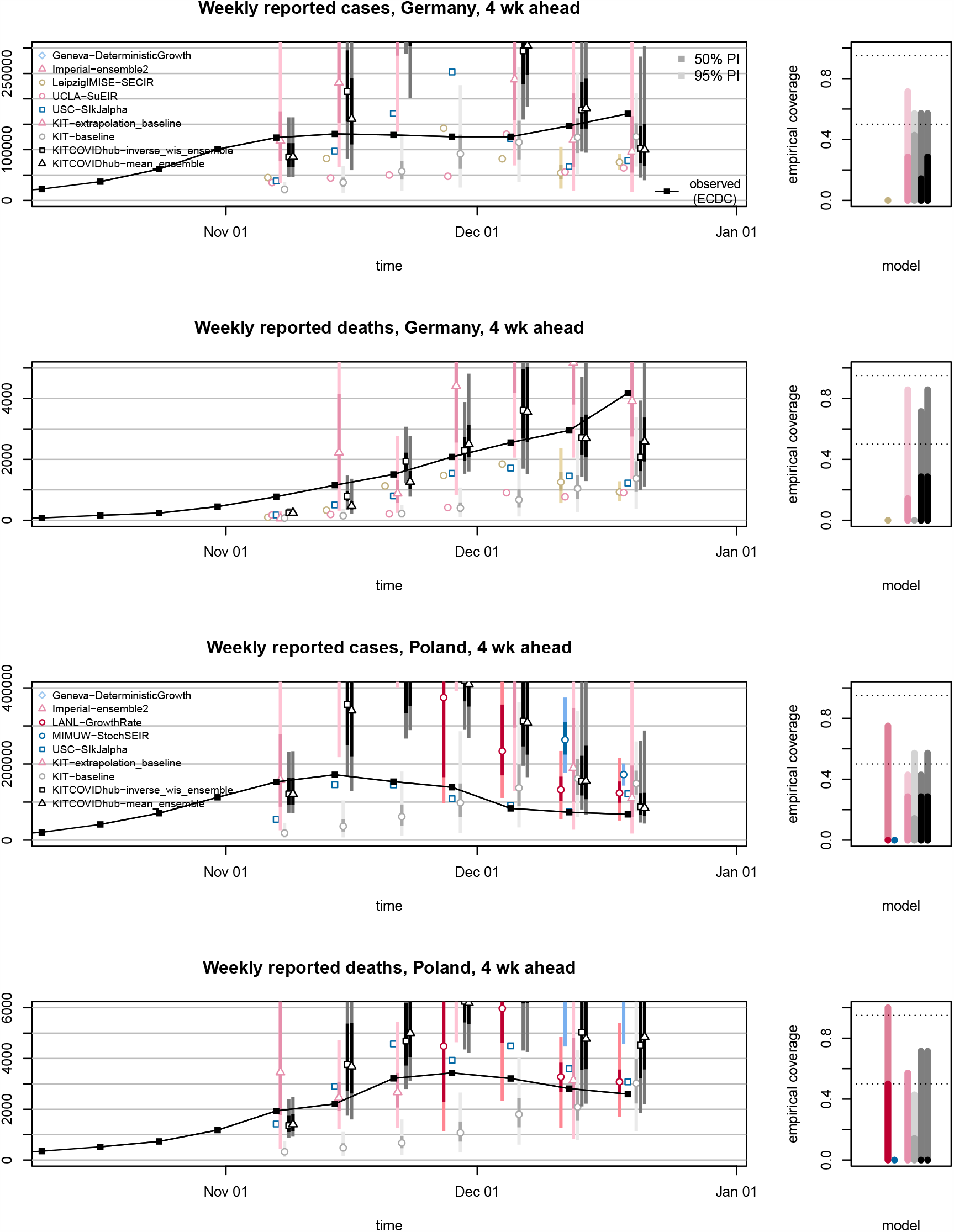
Four-week-ahead forecasts of incident cases and deaths in Germany and Poland (left). Displayed are predictive medians, 50% and 95% prediction intervals for models not shown in Figure 13. Coverage plots (right) show the empirical coverage of 95% (light) and 50% (dark) prediction intervals.

The median, mean and inverse-WIS ensemble showed overall good, but not outstanding relative performance in terms of mean WIS. Differences between the different ensemble approaches are relatively minor and do not indicate a clear ordering. We re-ran the ensembles retrospectively using all available forecasts, i.e. including those submitted late or excluded due to implausibilities. As can be seen from Supplementary Table 7, this led only to minor changes in performance. Unlike in the US effort (Brooks et al., 2020) the ensemble forecast is not strictly better than the single-model forecasts. Typically, performance is similar to some of the better-performing contributed forecasts, and sometimes the latter have a slight edge (e.g. `FIAS_FZJ-Epi1Ger` for cases in Germany and `MOCOS-agent1` for deaths in Poland). Interestingly, the expert forecast `epiforecasts-EpiExpert` is often among the more successful methods, indicating that an informed human assessment sets a high bar for more formalized model-based approaches. In terms of point forecasts, the extrapolation approach `Geneva-DetGrowth` shows good relative performance, but only covers one-week-ahead forecasts.

The 50% and 95% prediction intervals of most forecasts did not achieve their respective nominal coverage levels (most apparent for cases two weeks ahead). The statistical time series model `KIT-time_series_baseline` features favourably here, though at the expense of wide forecast intervals (Figure 3). While its lack of sharpness leads to mediocre overall performance in terms of the WIS, the model seems to have been a helpful addition to the ensemble by counterbalancing the overconfidence of other models. Indeed, coverage of the 95% intervals of the ensemble is above average, despite not reaching nominal levels.

A last aspect worth mentioning concerns the discrepancies between results for one-week-ahead incident and cumulative quantities. In principle these two should be identical, as forecasts should only be shifted by an additive constant (the last observed cumulative number). This, however, was not the case for all submitted forecasts, and coherence was not enforced by our submission system. For the ensemble forecasts the discrepancies are largely due to the fact that the included models are not always the same.

## 5 Conclusions

We presented results from a preregistered forecasting project in Germany and Poland, covering 10 weeks during the second wave of the COVID-19 pandemic. We believe that such an effort is helpful to put the outputs from single models in context, and to give a more complete picture of the associated uncertainties. For modelling teams, short-term forecasts can provide a useful feedback loop, via a set of comparable outputs from other models, and regular independent evaluation. A substantial strength of our study is that it took place in the framework of a prespecified evaluation protocol. The criteria for evaluation were communicated in advance, and most considered models covered the entire study period.

Similarly to Funk et al. (2020), we conclude that achieving good predictive accuracy and calibration is challenging in a dynamic epidemic situation. Part of the reason may be that not all models were designed for the sole purpose of short-term forecasting, and could be tailored more specifically to this task. Certain models were originally conceived for what-if projections and retrospective assessments of longer-term dynamics and interventions. This focus on a global fit may limit their flexibility to align closely with the most recent data, making them less successful at short forecast horizons compared to simpler extrapolation approaches. We observed pronounced heterogeneity between the different forecasts, with a general tendency to overconfident forecasting, i.e. too narrow prediction intervals. While over the course of ten weeks, some models showed better average performance (in terms of formal evaluation criteria) than others, relative performance has been fluctuating considerably. Different models may in fact be particularly suitable for different phases of an epidemic (Funk et al., 2020), which is exemplified by the fact that some models were quicker to adjust to slowing growth of cases in Germany. These aspects highlight the importance of considering several independently run models rather than focusing attention on a single one, as is sometimes the case in public discussions. Here, collaborative forecasting projects can provide valuable insights. Overall, ensemble methods showed good, but not outstanding relative performance, notably with clearly above-average coverage rates. Its improved reliability is a key strength of the ensemble approach, and we expect that the continuing refinement of member models will further strengthen the robustness of the ensemble. An important question is whether ensemble forecasts could be improved by sensible weighting of members or post-processing steps. Given the limited amount of available forecast history and rapid changes in the epidemic situation, this is a challenging encounter, and indeed we did not find benefits in the inverse-WIS approach.

An obvious extension to both assess forecasts in more detail and make them more relevant to decision makers is to issue them at a finer geographical resolution. During the evaluation period covered in this work, only three of the contributed forecast models (`ITWW-county_repro` and `USC-SIkJalpha`, `LeipzigIMISE-SECIR` for the state of Saxony) also provided forecasts at the regional level (German states, Polish voivodeships). Extending this to a larger number of models is one of the main priorities for the further course of the German and Polish Forecast Hub project.

In its present form, the project covers only forecasts of confirmed cases and deaths. These commonly addressed forecasting targets were already covered by a critical mass of teams when the project was started. Given limited available time resources of teams, a choice was made to focus efforts on this narrow set of targets. An extension to other quantities such as hospitalizations or ICU/ventilation need, which have important public health implications, was considered, but in view of emerging parallel efforts and open questions on data availability not prioritized.

The German and Polish Forecast Hub will continue to compile short-term forecasts and process them into forecast ensembles. With vaccine rollout likely to start in early 2021, models will face a new layer of complexity. We aim to provide further systematic evaluations for these future phases, contributing to a growing body of evidence on the potential and limits of pandemic short-term forecasting.

## Reproducibility / data availability

All data used in this article are publicly available at https://github.com/KITmetricslab/covid19-forecast-hub-de. Forecasts can be visualized interactively at https://github.com/KITmetricslab/covid19-forecast-hub-de. Codes to reproduce figures and tables are available at https://github.com/KITmetricslab/analyses_de_pl.

## Acknowledgements

We are grateful to the team of the US COVID-19 Forecast Hub, in particular Evan L. Ray and Nicholas G. Reich, for fruitful exchange and their support. We would like to thank Dean Karlen for contributions to the Forecast Hub from December 2020 onwards and Berit Lange for helpful comments. We moreover want to thank Fabian Eckelmann and Knut Persecke who implemented the interactive visualization tool.

The work of Johannes Bracher was supported by the Helmholtz Foundation via the SIMCARD Information and Data Science Pilot Project. Sangeeta Bhatia acknowledges support from the Wellcome Trust (219415). Nikos I. Bosse acknowledges funding by the Health Protection Research Unit (grant code NIHR200908). Sebastian Funk and Sam Abbott acknowledge support from the Wellcome Trust (grant no. 210758/Z/18/Z). The work of Ajitesh Srivastava was supported by National Science Foundation Award No. 2027007 (RAPID). Tilmann Gneiting and Daniel Wolffram are grateful for support by the Klaus Tschira Foundation.

The content is solely the responsibility of the authors and does not necessarily represent the official views of the institutions they are affiliated with.

## Author contributions

JB, TG and MS conceived the study with advice from AU. JB, DW, JD, KG and JK put in place and maintained the forecast submission and processing system. AU coordinated the creation of an interactive visualization tool. JB performed the evaluation analyses with inputs from DW, TG, MS and members of various teams. SA, MVB, DB, SB, MB, NIB, JPB, LC, GF, JF, SF, KG, QG, SH, TH, YK, HK, TK, EK, MLL, JHM, IJM, KN, TO, FR, MS, SS, AS, JZ and DZ contributed forecasts (see list of contributors by team). JB, TG and MS wrote the manuscript, with TK, MB and KG contributing to the description of intervention measures in Poland. All teams and members of the coordinating team provided feedback on the manuscript and descriptions of the respective models.

## List of contributors by team

`CovidAnalytics-DELPHI` Michael Lingzhi Li (Operations Research Center, Massachusetts Institute of Technology, Cambridr, MA, USA), Dimitris Bertsimas, Hamza Tazi Bouardi, Omar Skali Lami, Saksham Soni (all Sloan School of Management, Massachusetts Institute of Technology, USA)

`epiforecasts-EpiExpert` and `epiforecasts-EpiNow2` Sam Abbott, Nikos I. Bosse, Sebastian Funk (all London School of Hygiene and Tropical Medicine, London, UK)

`FIAS_FZJ-Epi1Ger` Maria Vittoria Barbarossa (Frankfurt Institute for Advanced Studies, Frankfurt, Germany), Jan Fuhrmann (Jülich Supercomputing Centre, Forschungszentrum Jülich, Jülich, Germany and Frankfurt Institute for Advanced Studies, Frankfurt, Germany), Jan H. Meinke (Jülich Supercomputing Centre, Forschungszentrum Jülich, Jülich, Germany)

`Geneva-DeterministicGrowth` Antoine Flahault, Elisa Manetti (both Institute of Global Health, Faculty of Medicine, University of Geneva, Geneva, Switzerland), Christine Choirat, Benjamin Bejar Haro, Ekaterina Krymova, Gavin Lee, Guillaume Obozinski, Tao Sun (all Swiss Data Science Center, ETH Zurich and EPFL Lausanne, Switzerland), Dorina Thanou (Center for Intelligent Systems, EPFL, Lausanne Switzerland)

`ICM-agentModel` L ukasz Górski, Magdalena Gruziel-Slomka, Artur Kaczorek, Antoni Moszyński, Karol Niedzielewski, Jedrzej Nowosielski, Maciej Radwan, Franciszek Rakowski, Marcin Semeniuk, Jakub Zieliński (all Interdisciplinary Centre for Mathematical and Computational Modelling, University of Warsaw, Warsaw, Poland), Rafal Bartczuk (Interdisciplinary Centre for Mathematical and Computational Modelling, University of Warsaw, Warsaw and Institute of Psychology, John Paul II Catholic University of Lublin, Lublin, Poland), Jan Kisielewski (Interdisciplinary Centre for Mathematical and Computational Modelling, University of Warsaw, Warsaw and Faculty of Physics, University of Bialystok, Ciolkowskiego 1L, 15-245 Bialystok)

`Imperial-ensemble2` Sangeeta Bhatia (MRC Centre for Global Infectious Disease Analysis, Abdul Latif Jameel Institute for Disease and Emergency Analytics (J-IDEA), Imperial College, London, UK)

`ITWW-county_repro` Przemyslaw Biecek (Warsaw University of Technology, Warsaw, Poland), Viktor Bezborodov, Marcin Bodych, Tyll Krueger (all Wroclaw University of Science and Technology, Poland), Jan Pablo Burgard (Economic and Social Statistics Department, University of Trier, Germany), Stefan Heyder, Thomas Hotz (both Institute of Mathematics, Technische Universität Ilmenau, Germany)

`LANL-GrowthRate` Dave A. Osthus, Isaac J. Michaud (both Statistical Sciences Group, Los Alamos National Laboratory, Los Alamos, USA), Lauren Castro, Geoffrey Fairchild (both Information Systems and Modeling, Los Alamos National Laboratory, Los Alamos, USA)

`LeipzigIMISE-SECIR` Yuri Kheifetz, Holger Kirsten, Markus Scholz (all Institute for Medical Informatics, Statistics and Epidemiology, University of Leipzig, Leipzig, Germany)

`MIMUW-StochSEIR` Anna Gambin, Krzysztof Gogolewski, Blażej Miasojedow, Ewa Szczurek (all Institute of Informatics, University of Warsaw, Warsaw, Poland), Daniel Rabczenko, Magdalena Rosińska (Polish National Institute of Public Health – National Institute of Hygiene)

`MOCOS-agent1` Marek Bawiec, Viktor Bezborodov, Marcin Bodych, Tyll Krueger, Tomasz Ożański, Barbara Pabjan, Ewaryst Rafajllowicz, Ewa Skubalska-Rafajlowicz, Wojciech Rafajllowicz (all Wroclaw University of Science and Technology, Poland), Przemyslaw Biecek (Warsaw University of Technology), Agata Migalska (Wroclaw University of Science and Technology, Poland and Nokia Solutions and Networks, Wroclaw, Poland), Ewa Szczurek (University of Warsaw)

`UCLA-SuEIR` Quanquan Gu, Pan Xu, Jinghui Chen, Lingxiao Wang, Difan Zou, Weitong Zhang (all Department of Computer Science, University of California, Los Angeles, USA)

`USC-SikJalpha` Ajitesh Srivastava, Viktor K. Prasanna, Frost Tianjian Xu (all University of Southern California, Los Angeles, USA)

## Supplementary Materials

### A Stratified visualizations of case and death counts

### B Detailed description of baseline forecasts

We here describe the three baseline forecasts from Section 3.1 in more detail.

#### B.1 KIT-baseline

Denote the quantity of interest on the incidence scale by *X*_*t*_. The corresponding quantity on the cumulative scale is denoted by *Y*_*t*_ = Σ_*s≤t*_ *X*_*t*_. The one-week-ahead forecast for *X*_*t*+1_ is given by a negative binomial distribution with mean *X*_*t*_ and overdispersion parameter *ψ*. Due to the skewness of the negative binomial distribution this implies that the predictive median is slightly smaller than *X*_*t*_. The overdispersion parameter is estimated from the last five available observations using a maximum likelihood approach, i.e. by maximizing

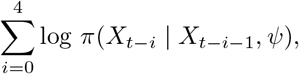

with respect to *ψ*, where *π*(*·* | *X*_*t−i−*1_, *ψ*) is the probability mass function of a negative binomial distribution with mean *X*_*t−i−*1_ and overdispersion parameter *ψ*. For technical reasons we replace any mean of a negative binomial distribution which would equal zero by 0.2. The two- to four-week-ahead forecasts are simply set to the same distribution as the one-week-ahead forecast.

To obtain forecasts on the cumulative scale we assume independence between *X*_*t*+1_, *X*_*t*+2_, *X*_*t*+3_ and *X*_*t*+4_. As the sum of independent random variables following negative binomial distributions with the same overdispersion parameter follows again a negative binomial distribution, *Y*_*t*+1_, *Y*_*t*+2_, *Y*_*t*+3_ and *Y*_*t*+4_ follow shifted negative binomial distributions with overdispersion parameter *ψ*, 2*ψ*, 3*ψ* and 4*ψ*, respectively.

### B.2 KIT-extrapolation baseline

We assume again a (conditional) negative binomial distribution, but with mean *λ*_*t*+1_ = *αX*_*t*_ rather than just *X*_*t*_. The parameter *α* is estimated from the last three observed values in the following way:

- If the last three observations are ordered, i.e. *X*_*t−*2_ *< X*_*t−*1_ *< X*_*t*_ or *X*_*t−*2_ *> X*_*t−*1_ *> X*_*t*_ we let

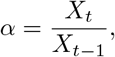

which corresponds to simple multiplicative extrapolation.
- Otherwise we let *α* = 1, so that the predictive mean *λ*_*t*+1_ equals the last observation *X*_*t*_.

The idea behind this distinction is that the model should only use trends if they have manifested for at least two weeks. The overdispersion parameter is estimated by maximizing

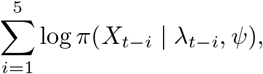

with respect to *ψ* (keeping the value *α* entering into *λ*_*t−i*_ = *αX*_*t−i−*1_ constant at the value chosen as described above). Note that we do not use the last observation *X*_*t*_ here as by construction (if the last three observations are ordered) *X*_*t*_ = *λ*_*t*_.

We then sample 100,000 paths (*X*_*t*+1_, *X*_*t*+2_, *X*_*t*+3_, *X*_*t*+4_) from this model and obtain forecast quantiles for both incident and cumulative quantities from these samples.

### B.3 KIT-time series baseline

We fit an exponential smoothing model with multiplicative errors and without seasonality to the last 12 observations on the incidence scale. The `R` (R Core Team, 2020) command is `forecast::ets(ts, model=“MMN”)` using the `forecast` package (Hyndman and Khandakar, 2008). As noted in the main text, this specification is taken from Petropoulos and Makridakis (2020). As in the previous section we proceed by sampling paths from this model and computing predictive quantiles from them.

### C Sources on changes in non-pharmaceutical interventions and testing regimes

We here provide sources for the dates of interventions shown in Figure 1.

#### Poland

Government interventions are largely documented on the respective governmental web site and the Twitter channel of the Polish Ministry of Health (in Polish):

- https://www.gov.pl/web/koronawirus/100-dni-solidarnosci-w-walce-z-covid-19
- https://twitter.com/MZ_GOV_PL.

Specific news items on mentioned interventions/events:

- Four symptoms required for test: Ministerstwo Zdrowia przekazalo zasady zlecania testów na koronawirusa, Wprost, 23 September 2020, https://www.wprost.pl/koronawirus-w-polsce/10368723/ministerstwo-zdrowia-przekazalo-zasady-zlecania-testow-na-koronawirusa.html (last accessed 22 December 2020).
- Only one out of four symptoms required for test: Dlaczego lekarz odmawia skierowania na test na COVID-19? Medonet, 5 Nov 2020, https://www.medonet.pl/koronawirus/koronawirus-w-polsce,kiedy-lekarz-moze-odmowic-skierowania-na-test-na-koronawirusa,artykul,26303647.html (last accessed 22 December 2020)
- Bulk reporting of 22,000 cases on 25 November: Rozbiezności w statystykach koronawirusa. 22 tys. przypadków będą doliczone do ogólnej liczby wyników, Forsal, 23 November, https://forsal.pl/lifestyle/zdrowie/artykuly/8017628,rozbieznosci-w-statystykach-koronawirusa-22-tys-przypadkow-beda-doliczone-do-ogolnej-liczby-wynikow.html (last accessed 22 December 2020)
- High test positivity and suspected under-ascertainment: Polish doctors fear high rate of positive COVID tests show pandemic worse than it appears, J. Plucinska, Reuters, 1 December 2020, https://www.reuters.com/article/us-health-coronavirus-poland-cases/polish-doctors-fear-high-rate-of-positive-covid-tests-show-pandemic-worse-than-it-appears-idUSKBN28B54Q (last accessed 22 December 2020)

#### Germany

- A chronicle of the most important events (in German) can be found on the web site of the Germany Ministry of Health:
- https://www.bundesgesundheitsministerium.de/coronavirus/chronik-coronavirus.html

Specific news items on mentioned interventions/events:

- New testing strategy announced: SARS-CoV-2-Diagnostik: RKI passt Testempfehlungen an, Ä rzteblatt, 3 November 2020, https://www.aerzteblatt.de/nachrichten/118001/SARS-CoV-2-Diagnostik-RKI-passt-Testempfehlungen-an (last accessed 22 December 2020)
- Semi-lockdown from 2 November onwards: Coronavirus: Germany to impose one-month partial lock-down, Deutsche Welle, 28 October 2020, https://www.dw.com/en/coronavirus-germany-to-impose-one-month-partial-lockdown/a-55421241 (last accessed 22 December 2020)
- Reinforced rules from 1 December onwards: Was gilt wo im Corona-Dezember? Tagesschau, 1 December 2020, https://www.tagesschau.de/inland/corona-plan-bundeslaender-beschluss-103.html (last accessed 22 December 2020)
- Full lockdown starting on 16 December: Lockdown in Deutschland – Das sind die Corona-Regeln. Tagesschau, 13 December 2020, https://www.tagesschau.de/inland/corona-regeln-lockdown-101.html (last accessed 22 December 2020)

### D Availability and delays of forecasts

Each entry describes up to which forecast horizon (in weeks) forecasts for incident cases, cumulative cases, incident deaths and cumulative deaths were made available (numbers in this order and separated by semicolons). Asterisks indicate that forecasts were only available on Wednesday or later rather than before Tuesday 3pm.

### E Additional results for one- and two-week-ahead forecasts

### F Results for three- and four-week-ahead forecasts

